# Addressing Label Noise for Electronic Health Records: Insights from Computer Vision for Tabular Data

**DOI:** 10.1101/2023.10.17.23297136

**Authors:** Jenny Yang, Hagen Triendl, Andrew A. S. Soltan, Mangal Prakash, David A. Clifton

## Abstract

The analysis of extensive electronic health records (EHR) datasets often calls for automated solutions, with machine learning (ML) techniques, including deep learning (DL), taking a lead role. One common task involves categorizing EHR data into predefined groups. However, the vulnerability of EHRs to noise and errors stemming from data collection processes, as well as potential human labeling errors, poses a significant risk. This risk is particularly prominent during the training of DL models, where the possibility of overfitting to noisy labels can have serious repercussions in healthcare. Despite the well-documented existence of label noise in EHR data, few studies have tackled this challenge within the EHR domain. Our work addresses this gap by adapting computer vision (CV) algorithms to mitigate the impact of label noise in DL models trained on EHR data. Notably, it remains uncertain whether CV methods, when applied to the EHR domain, will prove effective, given the substantial divergence between the two domains. We present empirical evidence demonstrating that these methods, whether used individually or in combination, can substantially enhance model performance when applied to EHR data, especially in the presence of noisy/incorrect labels. We validate our methods and underscore their practical utility in real-world EHR data, specifically in the context of COVID-19 diagnosis. Our study highlights the effectiveness of CV methods in the EHR domain, making a valuable contribution to the advancement of healthcare analytics and research.

## 1 Introduction

In recent years, there has been a substantial surge in digital data within the healthcare sector. Notably, electronic health records (EHRs), which encompass patient health information such as medical history, diagnoses, medications, and lab test results, have played a pivotal role in enhancing patient safety, streamlining care coordination, and improving efficiency. The widespread adoption of EHRs has translated into a substantial increase in the volume of digital data, providing a robust foundation for machine learning (ML) applications in healthcare. Leveraging this wealth of data, ML models can enhance precision, develop personalized treatment strategies, and enable predictive analytics, ultimately resulting in improved patient outcomes and more efficient healthcare delivery.

However, while EHRs serve as a valuable data source for ML tasks, their utilization can pose challenges due to potential noise and errors. These issues can arise from various common sources, including data entry errors (4; 21; 41; 29), incomplete information (4; 21; 41), inconsistencies (21; 47; 17), system errors (18; 23; 30), and diagnostic test errors (40; 31; 11; 2).

Previous studies have highlighted the need for greater effort to improve the accuracy and completeness of EHR data (2; 16; 33; 15; 49). For example, a survey study published in 2020 asked a total of 136,815 patients at three US healthcare organizations to read their EHR notes and identify any errors (2). Of the 29,656 patients who provided a response, 1 in 5 reported a mistake, with 40% of these mistakes being perceived by the patient as serious. Among patient-reported serious errors, the most common mistakes included those related to diagnoses, medical history, medications, and test results. Another record-review study published in 2018 investigated the errors and causes of failure in the communication of patients’ information between different hospital information/EHR systems (16). Through the review of 882 hospital records, the study identified 1,256 errors of 41 different types. These errors were classified into system level errors (65%) and operator-dependent errors (35%), and further stratified into four categories: administrative-financial errors (61%), errors related to national codes (23%), clinical errors (9%), and other errors (7%). The presence of errors in EHR data can have serious consequences for patient care and outcomes, as well as for research and analysis that relies on this data. Therefore, ensuring the accuracy and completeness of EHR data is an ongoing challenge for healthcare providers and researchers.

Despite the substantial evidence pointing to the prevalence of noisy and erroneous EHR data, the existing ML models documented in the literature for EHR data analysis have yet to address this concern. These models typically operate under the assumption that the data and labels are free from unwanted noise and corruption, a premise that does not accurately reflect real-world datasets. Additionally, it is widely acknowledged that ML models (especially deep learning [DL] models), are susceptible to overfitting to noisy labels (38; 42; 55). This susceptibility can lead to significant consequences, such as reduced generalization performance on unseen patient EHR records, unreliable predictions, the perpetuation of undesired biases in predictions, and potentially serious repercussions for patient care. Consequently, this could erode trust among healthcare professionals regarding the utilization of ML/DL models. Therefore, it is crucial to acknowledge the inherent imperfect nature of EHR data and devise mechanisms for training ML/DL methods to effectively handle noisy data.

Expanding upon previous research, we acknowledge the presence of noisy data and labels during the training of DL models and aim to tackle this concern within the context of EHR data. Thus, our study is centered on EHR classification, with particular emphasis on situations where only the class labels are affected by noise or errors. Taking inspiration from recent methodologies in computer vision (CV) that account for noisy labels when training DL models (14; 54; 22), we undertake an exploration of their suitability for the EHR domain. It is essential to highlight that our tabular EHR data is distinctively different from image data for which these CV methods are originally proposed for. In contrast to images, where pixel values represent visual features, tabular EHR data encompasses a diverse array of patient records, diagnostic codes, timestamps, and various clinical parameters. Additionally, the amount of high quality data available in CV (with benchmarks like ImageNet (58) for training DL models) is typically many magnitudes larger than that available for EHRs. The differences between these two domains calls for ensuring that these methods function effectively and provide meaningful insights when applied to EHR.

In this study, we aim to bridge this gap. With relatively little adaptations tailored to the unique characteristics of EHR data, we found that some of the recently proposed methods in CV domain can substantially mitigate the risks associated with overfitting to noisy labels in EHR data. This finding highlights the adaptability and potential of these techniques, even in the presence of substantial differences in data structure and content. Moreover, our research goes beyond the individual application of these methods. We investigate the synergy of combining multiple approaches, and our results demonstrate that the integration of these techniques not only effectively addresses the issue of noisy labels, but also surpasses the performance of each method independently in many cases. Our findings emphasize the potential to transform how we handle and analyze EHR data, offering new avenues for improved healthcare outcomes and research in the EHR domain.

## 2 Related Works

Various techniques have been employed in different domains to address the challenges posed by noisy labels for ML tasks. In general, these can roughly be divided into two groups, 1) label correction, which focuses specifically on identifying and rectifying mislabeled data points to improve the quality of the training data, and 2) regularization, which penalises over-confident predictions to prevent overfitting and indirectly reduce the impact of noisy labels.

With respect to label correction, one approach is data cleaning (7), which involves removing data points that are clearly incorrect or inconsistent. This can be achieved through manual inspection or by clustering (26; 35) and outlier detection algorithms (19; 6; 3). However, manual inspection is expensive and time-consuming (7); and removing samples wastes valuable information that could still provide useful information for training (51). Conversely, algorithm-driven approaches, including self-training (43; 20; 12) and co-training (5; 25), iteratively update and improve labels based on a model’s predictions. These methods can be effective in improving the quality of labeled data; however, they typically rely on the initial labeled data. Thus, if the initial labels are noisy or biased, these can be propagated during training, potentially exacerbating the problem. Similarly, these techniques can lead to overconfident predictions on noisy data, especially if the model is uncertain about the true labels, resulting in the inclusion of incorrect labels in the training set. Additionally, these techniques may not be well-suited for very small datasets where the benefits of leveraging unlabeled data might be limited.

Given that these methods rely on the assumption of having access to a limited, dependable set of clean samples, our research will focus on situations where this assumption does not hold, which is frequently encountered in clinical data scenarios. Consequently, we will focus our investigations on regularization methods. These include robust loss functions, label smoothing, taking a convex combination of samples/labels, and using consistency as a metric for evaluation.

Robust loss functions can be utilized to minimize the impact of outliers and noisy labels in the loss function (10; 48). While employing these outlier-robust loss functions offer advantages, they also exhibit drawbacks, including the potential loss of crucial information, especially notable in real healthcare data, where outliers can yield valuable insights. The loss of outlier information can also introduce bias into a model’s predictions, potentially skewing them away from the true underlying data distribution. Furthermore, it’s important to acknowledge that within healthcare data, a spectrum of disease severity exists. Consequently, even when incorrect labels are present, the individual data samples themselves may not necessarily qualify as outliers.

Label smoothing (22; 39; 44) is often used in cases where data is imperfectly labeled or contains errors. It aims to improve the generalization and robustness of a model by preventing it from overfitting to the training data. It does this by adding small amounts of uncertainty to the target labels while the model is being trained, encouraging the model to assign lower probabilities to incorrect classes and distribute the probability mass more evenly across all classes. Label smoothing has previously been demonstrated to effectively remove noise from corrupted labels (22).

Mix-up (54) is a data augmentation technique commonly used in CV for tasks such as image classification. It was introduced as a regularization method to improve the generalization and robustness of models, especially in scenarios with limited labeled data. Combining different features and labels with one another prevents the model from becoming overconfident about the relationship between features and their labels, thereby regularizing the model.

Previous studies have also leveraged the concept of generating consistent outputs as a means to constrain training. One such method is bootstrapping (28), whereby the standard prediction objective is enhanced with a term for perceptual consistency. Here, a prediction is defined as consistent when a network produces the same prediction when presented with similar features. Another method is Neighbour Consistency Regularization (NCR) (14). Similarly, NCR enforces consistency among the predictions of a model on neighboring/similar samples. During training, the model is trained using both a standard supervised loss (such as cross-entropy) and an additional regularization term that encourages the predictions of neighboring samples to be consistent. By doing so, a model learns to be robust to label noise, as the regularization term penalizes inconsistencies in the predictions caused by noisy labels.

Although not specific to addressing noisy labels, ensemble methods offer another possible avenue for mitigating the effects of noisy labels. By training multiple models on different subsets of the data and combining their predictions, ensemble methods can help reduce the influence of individual noisy labels.

In the domain of EHR data analysis, limited studies to date address the problem of label noise. Very recently, Tjandra et al (57) tackled the problem of instance dependent label noise in EHR data where the authors assume the availability of a small subset of clean data and labels in addition to a larger noisy dataset for learning a two-stage discriminative model. Boughorbel et al (56) present an alternating loss-correction approach for training models with longitudinal EHR data with noisy labels. This method also requires the availability of separate clean and noisy datasets and alternates the training between these two sets. Unlike these approaches, we do not assume any prior knowledge about which datapoints are labeled correctly or incorrectly, and hence we do not require separate clean and noisy datasets. We show that even for EHR datasets, methods like NCR (14), Mix-up (54) and Label smoothing (22; 39; 44) (which do not need any additional information from a separate clean dataset) can be effectively applied with minor adaptations, thereby offering new avenues for improved EHR data analysis.

## 3 Data and Methods

We focus specifically on a real-world COVID-19 case study where the task is to use patients’ EHRs to classify them as being either COVID positive or negative. With government regulation for mandatory testing at the time, there is vast, clinically-rich EHR data available, alongside positive and negative COVID-19 presentations (determined through PCR tests, the gold standard test for diagnosing viral genome targets). Additionally, due to incomplete penetrance of PCR testing during the early stages of the pandemic and imperfect sensitivity, there is uncertainty in the viral status of patients who tested negative, thus making this case study uniquely suitable to test model development in the presence of noisy/incorrect labels.

### 3.1 Datasets and Pre-processing

For the purpose of this study, we use the CURIAL datasets (36; 37), which consist of anonymized EHR data for patients presenting to emergency departments (EDs) across four independent United Kingdom (UK) National Health Service (NHS) Trusts. These include Oxford University Hospitals NHS Foundation Trust (OUH), University Hospitals Birmingham NHS Trust (UHB), Portsmouth Hospitals University NHS Trust (PUH), and Bedfordshire Hospitals NHS Foundations Trust (BH). United Kingdom NHS approval via the national oversight/regulatory body, the Health Research Authority (HRA), has been granted for development and validation of artificial intelligence models to detect COVID-19 (CURIAL; NHS HRA IRAS ID: 281832).

Previous studies have shown that ML classification models trained on EHR features could diagnose patients presenting with COVID-19 up to 26% sooner than lateral flow device (LFD) testing and 90% sooner than PCR testing (36) on average, while simultaneously achieving high sensitivities and performing effectively as a rapid test-of-exclusion (36; 37; 50; 51; 52; 53). Similarly, we trained models for the purpose of rapid triage using laboratory blood tests and vital signs, as these are routinely collected during the first hour of patients attending emergency care in hospitals in middle- to high-income countries (37). The feature sets included are the same as those used in (37; 50; 51; 52; 53). Supplementary Section C summarizes the final features used.

For model development, a training set was used for model training; a validation set was used for continuous validation and threshold adjustment; and after successful development, internal and external test sets were used to evaluate the performance of final models. For training and validation, we used patient presentations from PUH. From PUH, we obtained patient presentations to the ED between March 1, 2020 and February 28, 2021. We created training, validation, and test sets using randomly selected 60%, 20%, and 20% splits, respectively. This resulted in 22,737 (1,182 COVID-19 positive), 7,579 (439 positive), and 7,580 (385 positive) presentations for PUH training, validation, and test sets, respectively.

We additionally curated datasets from three independent hospitals - OUH, UHB, and BH. From OUH, we curated two data extracts. The first extract contains 701 COVID-19 positive cases from the “first wave” of the COVID-19 epidemic in the UK (December 1, 2019 to June 30, 2020), with 91,970 pre-pandemic controls (COVID-free patient presentations from OUH prior to the global COVID-19 outbreak). The second extract contains 22,857 presentations (2,012 positive) from the “second wave” (October 1, 2020 – March 6, 2021). There was one cohort from UHB (presentations between December 1, 2019 and October 29, 2020) and BH (presentations between January 1, 2021 and March 31, 2021), consisting of 10,293 (439 positive) and 1,177 (144 positive), respectively. These cohorts were used to externally validate performance and generalizability, emulating the real-world implementation of such a diagnostic method.

Consistent with previous studies, we addressed the presence of missing values by using population median imputation, then standardized all features in our data to have a mean of 0 and a standard deviation of 1. A summary of the inclusion and exclusion criteria for patient cohorts, summary population statistics, and all training, validation, and test cohort splits can be found in Section C of the Supplementary Material.

For experiments with synthetic noise, we randomly changed the label to the incorrect diagnosis (i.e. increasing the false-negative and false-positive levels). For COVID-19 diagnosis by PCR, sensitivities were estimated to be around 80%-90% during different times of the pandemic (46; 24; 1), while specificity was estimated to be around 98%-100% (46; 24; 1). Thus, to represent more realistic label corruption, we created additional false negatives using 10%, 20%, 30%, and 40% of the COVID-19 positive presentations. Simultaneously, we kept the number of additional false positives consistent, representing 0.5% of the COVID-19 negative cases. This approach was chosen to a) mirror the estimated specificity of PCR testing for COVID-19 and b) address the significant label imbalances prevalent in the training data.

### 3.2 Baselines

As a baseline, we use the same general neural network architecture using cross-entropy loss shown to be successful in (50; 51; 52; 53) for the COVID-19 classification task (varying hyperparameters depending on the best results obtained during grid search). Additionally, (36; 37; 52; 53) showed that XGBoost works remarkably well for this task, and serves as another strong baseline.

#### 3.2.1 XGBoost

XGBoost (8) is an ensemble method which combines the predictions of several base estimators (in this case, decision trees) in order to improve generalizability and robustness. The idea is that the weaknesses of one model can be compensated for by the strengths of another, resulting in a more robust ensemble model.

#### 3.2.2 Baseline Neural Network

Following (50; 51; 52; 53), we trained a fully-connected neural network which used the rectified linear unit (ReLU) activation function in the hidden layers and the sigmoid activation function in the output layer. For updating model weights, the Adam optimizer was used during training. Details of the architecture are presented in Section B of the Supplementary Material. This architecture will also be referred as *Baseline NN* going forward.

### 3.3 CV-inspired Techniques to Address Noisy Labels

Here we briefly describe three recently proposed methods which were shown to be effective for training neural networks with noisy labels in CV tasks. We investigated both their individual and combined effectiveness in mitigating the impact of label noise in EHRs when employed in conjunction with the *baseline NN* described in 3.2.2.

#### 3.3.1 Label Smoothing

Label smoothing (39; 44; 22) is a regularization technique that adds a small amount of noise to the target labels during training. Similar to (22; 44), instead of using 0 or 1 as the correct label, we use a value of 1 − *ϵ* for the correct label and 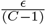 for the other labels. Here, C is the number of classes, and *ϵ* is sampled uniformly in [0, 1].

#### 3.3.2 Mix-Up

Mix-up(54) is an augmentation method that creates new examples as convex combinations of the original training samples. Given a dataset with labeled examples, Mix-up combines pairs of input samples (both the features and labels) by taking a weighted linear combination.

Therefore, for two data points (*x*_1_, *y*_1_) and (*x*_2_, *y*_2_), the mixed data point (*x*_*mix*_, *y*_*mix*_) is computed as follows:

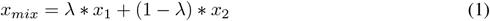

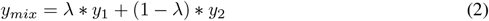

Here, *λ* denotes a mixing coefficient, which is sampled from a beta distribution, *Beta*(*α, α*), where *α* is a hyperparameter controlling the shape of the distribution, and *α* ∈ (0, ∞) (54). In our implementation, we also set a probability of mix-up per batch.

#### 3.3.3 Neighbour Consistency Regularization

Neighbour Consistency Regularization (NCR) (14) is a regularization technique which relies on enforcing the simple idea that examples from the same class will have similar latent representations and hence should be classified to the same class irrespective of their labels (which may be noisy and different). As presented in (14), we define the similarity between two examples by the cosine similarity of their feature representations:

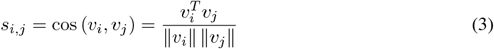

Here, the feature representations are non-negative values (obtained after a ReLU transformation) from a specific layer hidden layer. If *v*_*i*_ and *v*_*j*_ have high cosine similarity *s*_*i,j*_, then a classifier *f*, is encouraged to predict the same label for *f*(*v*_*i*_) and *f*(*v*_*j*_), regardless of their labels *y*_*i*_ and *y*_*j*_. This discourages the model from overfitting to any incorrect mapping (*x, y*), if either (or both) of *y*_*i*_ and *y*_*j*_ are noisy.

To enforce neighbor consistency regularization, the objective function is formulated to minimize the distance between logits **z**_**i**_ and **z**_**j**_, when their corresponding feature representations *v*_*i*_ and *v*_*j*_ are similar. Using 3, the NCR term can be written as:

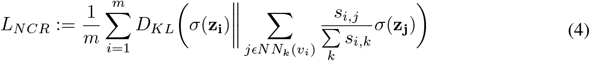

In this formulation, the *D*_*KL*_ represents the Kullback-Leibler (KL) divergence loss used to measure the dissimilarity between two distributions. The term *NN*_*k*_(*v*_*i*_) refers to the set of *k* nearest neighbors of *v*_*i*_ in the feature space. To ensure that the similarity values form a probability distribution, we normalize them. Additionally, we set the self-similarity *s*_*i,i*_ to zero to avoid it dominating the normalized similarity. Gradients are propagated back to all inputs. Thus, this NCR term encourages the output of a classifier to classify *x*_*i*_ in a way which aligns to its latent space neighbors, regardless of the potentially noisy label *y*_*i*_.

We combine this NCR with the standard supervised classification loss function, namely cross entropy, to form the final objective function that is minimized during training, i.e.,

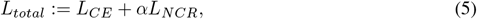

Here, the hyper-parameter *α* controls the strength of the NCR term. This differs slightly from the implementation in (14), where the authors vary the NCR term by *α* and the CE term by 1 − *α* (however, both implementations adjust the relative contributions of the CE and NCR loss terms). As alternatives to KL-divergence, we also investigate the effect of Jensen–Shannon divergence and mean absolute error within the NCR term. Results for these additional metrics can be found in Section F of the Supplementary Material.

### 3.4 Evaluation Metrics

We evaluate the trained models with commonly used classification metrics: area under the receiver operator characteristic curve (AUROC), area under the precision recall curve (AUPRC), sensitivity and specificity, alongside 95% confidence intervals (CIs) based on 1,000 bootstrapped samples taken from the test set. Tests of significance (represented by p-values), comparing the accuracies between models, are calculated by evaluating how many times one model performs better than another, across 1, 000 pairs of bootstrapped iterations drawn from the test set.

### 3.5 Hyperparameter Optimization and Threshold Adjustment

Hyperparameter values were chosen through grid search and standard five-fold cross-validation, using the training set (note that the validation and held-out test sets are used in threshold adjustment and final model evaluation, respectively). Grid search was used to determine the number of hidden layers of *baseline NN*, the number of nodes used in each layer, the learning rate, the max depth of XGBoost, the number of nearest neighbours in NCR, the weight of the NCR term, the starting epoch for NCR, and the ϵ and λ values used in Label Smoothing and Mix-up, respectively. Details on the hyperparameter values used in the reported results can be found in Supplementary Table 3.

In ML classification models, the output typically represents the probability of the input belonging to a certain class, where often a threshold on this probability needs to be set to determine a discrete label. We determined this threshold via grid search on the validation split.

For our specific objective, we tuned the threshold to achieve sensitivities of 0.85, ensuring that the model maintains clinically acceptable performance in effectively identifying positive COVID-19 cases. This sensitivity was chosen to exceed lateral flow device (LFD) tests, which achieved a sensitivity of 56.9% (95% confidence interval 51.7%-62.0%) for OUH admissions between December 23, 2021 and March 6, 2021 (36). Additionally, the gold standard for diagnosing viral genome targets is by real-time PCR (RT-PCR), which had estimated sensitivities around 80%-90% during different times of the pandemic (46; 24; 1).

## 4 Results

### 4.1 Comparison of Methods

In Figure 1, we compare the performance of all the methods in terms of AUROC for different amounts of label noise. Full numerical results can be found in Supplementary Table 6. The regularizers from CV domain provide significant improvement over the baselines with Mix-up and NCR emerging as the best performing methods in 14 out of 25 test sets. Since both the Mix-up and NCR methods demonstrated strong performance, we also conducted an assessment of their combined use. The model performances remained fairly consistent across the Mix-up, NCR, and combined Mix-up and NCR methods, with the combined method consistently outperforming the others. NCR or Mix-up method followed as the second-best performers in 18 out of 25 test cases. However, the difference in accuracy between Mix-up (both independently and when combined with NCR) compared to NCR, was not found to be significant (*p>* 0.05, based on 1,000 bootstrapped iterations; exact p-values can be found in the Supplementary Table 5). Amongst the CV methods, label smoothing performed the worst, steadily decreasing in AUROC as the error in the training data increased, across all test sets (AUROC decreased by up to 3% when there was 40% error in COVID-19 positive cases). A more detailed discussion of these results can be found in Section 5.

**Figure 1:**
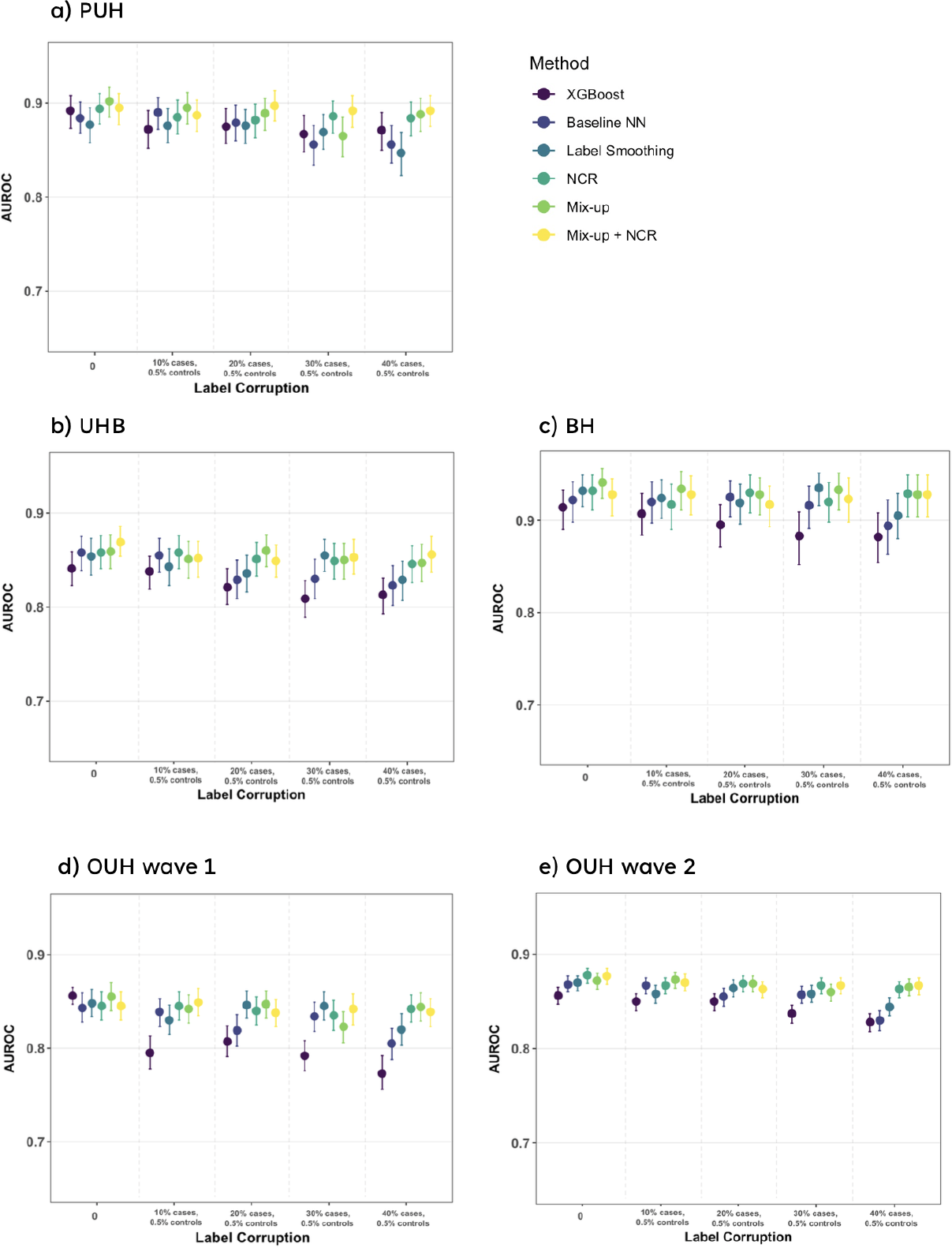
Change in performance (AUROC) at different training label corruption levels across different methods. Panels a)-e) show results for different test sets.

When considering the mean AUROC performances (alongside standard deviation) for each test set (see Supplementary Table 4), across different methods, we found that models trained with NCR (both alone and combined with Mix-up) exhibited lower standard deviations, indicating more consistent classification performance across different noise ratios. Standard deviations were between 0.004-0.008 for NCR and Mix-up+NCR methods, across all test sets, compared to standard devisions of *>*0.010 for all other methods (except Mix-up, where standard deviation was 0.005-0.006 for three of the five test sets). This implies that the performance drop with increasing noise was reduced with NCR compared to other methods. This is true across all test sets, suggesting that the models trained with NCR are robust and generalizable across independent and unseen cohort distributions. Full numerical values can be found in Supplementary Table 4.

Motivated by these findings, we conducted a more comprehensive assessment of training with NCR. With respect to clinical applications, we chose to focus on NCR since it does not involve altering the data. This is in contrast to Mix-up, which might be considered as using “synthetic data,” and may potentially be unsuitable for clinical tasks. NCR also performed closely to that of Mix-up and the combination of Mix-up and NCR. These results fell within confidence intervals, with accuracies that exhibited no significant differences across 1,000 bootstrapped runs.

### 4.2 Extended Analysis with Neighbour Consistency Regularization

After performing grid search, using five-fold standard cross validation, we determined the best hyperparameters to use in training (final hyperparameter values used can be found in Supplementary Table 3. The following results shown are for NCR with KL-divergence as mentioned in Equation 4. This is the same formulation as presented in the original NCR paper (14); however, we additionally performed similar analyses with other divergence measures. Results for these have been reported in Section F of the Supplementary Material. Our results indicate that these divergence measures produce outcomes similar to those achieved when utilizing KL-divergence.

#### 4.2.1 Ablation Study

We further investigate the effect of the key hyperparameters of NCR at different noise levels. Specifically, we analyze the impact of the hidden layer used for extracting the latent representations for calculating the NCR term, the weight of the NCR term, the epoch at which NCR is initialized, the batch size used, and the number of neighbors *k*. A subset of results for only PUH and BH datasets is shown in Fig. 2 for different noise levels and the full set of results (including the hidden layer and the batch size used) can be found in Supplementary Fig. 4. Fig. 2 illustrates that increasing the weight of NCR can improve performance at higher noise ratios, up to a certain extent. Furthermore, it is clear that irrespective of the noise level, providing a *warm-up* period of around 30 epochs with just the binary cross-entropy loss enabled (before switching on the NCR loss term) helps achieve superior performance than enabling both loss terms (as shown in Equation 4) from the start of training. A detailed discussion of these results can be found in Section 5. Finally, we also observe that considering around 10 nearest neighbors in the NCR loss formulation achieves the best results, with performance saturating with increasing number of neighbors. Hence, we ran all our experiments with 10 neighbors. Complete ablation results including all test sets and hyperparameters, across different noise ratios, can be found Supplementary Figure E).

**Figure 2:**
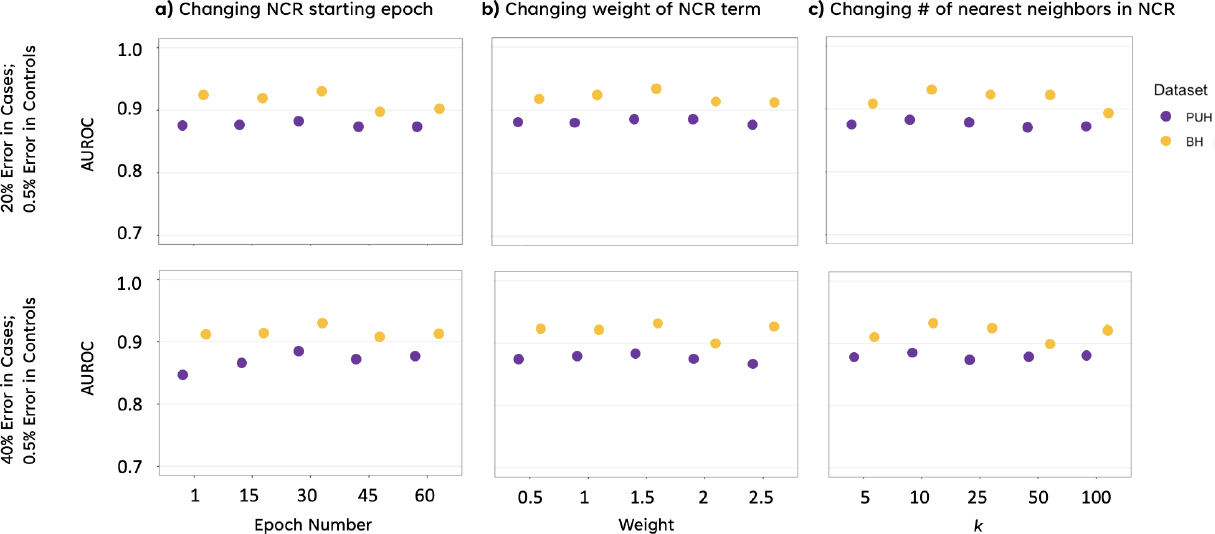
**Ablation study showing model performance with NCR with varying NCR hyperparameters for PUH and BH test sets**, across varying a) NCR starting epochs, b) NCR weights, and c) number of nearest neighbors (k). Results presented for 20% error in cases and 0.5% error in controls, and 40% error in cases and 0.5% error in controls.

**Figure 3:**
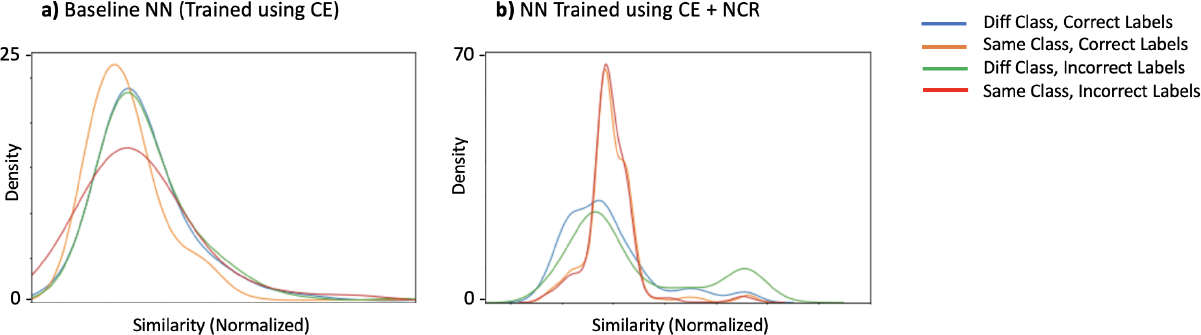
Feature similarity distributions at the end of training, for samples correctly and incorrectly labelled, across both similar and different classes without and with NCR. Feature similarity is calculated using cosine similarity as described in Equation 3. Results shown are for models trained on data with 40% noisy labels.

**Figure 4:**
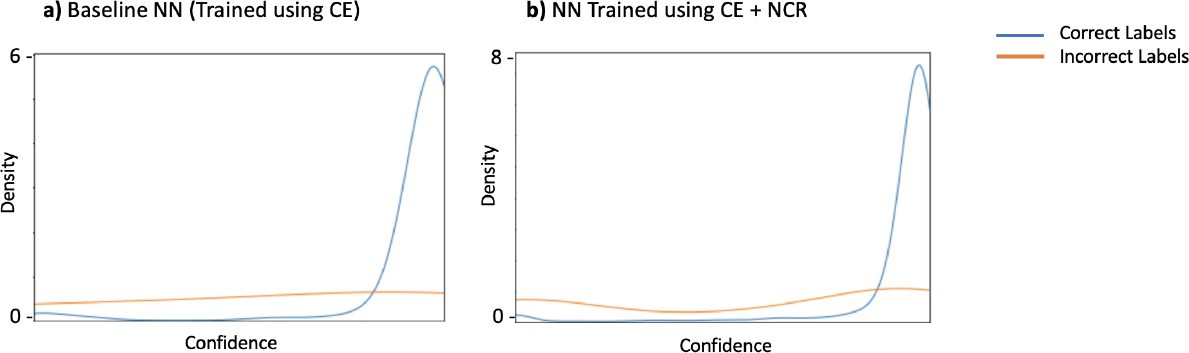
Predicted confidence of correctly labeled and incorrectly labeled training examples during different stages of training. Panel a) shows the predicted confidences when using Baseline NN trained using only cross entropy (CE) loss and b) that of the same model when trained using CE+NCR loss terms.

In comparison to the *baseline NN* model, NCR achieves a significant improvement in performance, reaching up to a 3.7% increase across all noise ratios for all test sets (Table 1). Surprisingly, the use of NCR achieves performance that is comparable, and in some cases even superior, to that of standard cross entropy when there is 0% added noise (*p* > 0.05), suggesting a general regularization effect of NCR.

**Table 1:** AUROC, AUPRC, Sensitivity, and Specificity comparison between baseline and NCR models, across different amounts of error and test sets. In addition to label error in COVID-19 positive cases, there is also 0.5% label error in the negative controls. 0% error represents the original dataset, without any added label noise. p-values shown compare differences in performance between the baseline NN (trained with cross entropy loss only) and the same model trained with NCR in addition to cross entropy loss (denoted by NCR). Bold faced values for each metric denote best performing method across different test sets.

| Test Set | AUROC |  | AUPRC |  | Sensitivity |  | Specificity |  | p-value |
| --- | --- | --- | --- | --- | --- | --- | --- | --- | --- |
|  | Baseline NN | CE+NCR | Baseline NN | CE+NCR | Baseline NN | CE+NCR | Baseline NN | CE+NCR |  |
| 0% error |  |  |  |  |  |  |  |  |  |
| PUH | 0.884(0.868-0.901) | **0.894(0.878-0.910)** | 0.538(0.494-0.583) | **0.579(0.536-0.624)** | **0.841(0.81-0.871)** | 0.807(0.774-0.838) | 0.722(0.713-0.731) | **0.831(0.824-0.838)** | p=0.38 |
| UHB | 0.858(0.839-0.875) | 0.858(0.841-0.876) | 0.309(0.278-0.340) | **0.387(0.348-0.431)** | **0.861(0.833-0.889)** | 0.831(0.800-0.859) | 0.638(0.629-0.646) | **0.723(0.716-0.731)** | p=0.396 |
| BH | 0.922(0.898-0.942) | **0.932(0.911-0.949)** | 0.691(0.627-0.751) | 0.753(0.690-0.816) | 0.931(0.894-0.961) | 0.903(0.859-0.941) | 0.659(0.636-0.684) | **0.770(0.748-0.790)** | p=0.346 |
| OUH "wave 2" | 0.868(0.860-0.877) | **0.878(0.869-0.885)** | 0.554(0.536-0.573) | **0.635(0.616-0.653)** | **0.875(0.862-0.887)** | 0.845(0.831-0.858) | 0.604(0.598-0.609) | **0.717(0.712-0.722)** | p=0.485 |
| OUH "wave 1" | 0.843(0.828-0.859) | **0.845(0.830-0.860)** | 0.087(0.078-0.097) | **0.155(0.133-0.181)** | **0.859(0.837-0.882)** | 0.806(0.78-0.83) | 0.615(0.613-0.618) | **0.729(0.726-0.731)** | p=0.273 |
| Error in 10% cases |  |  |  |  |  |  |  |  |  |
| PUH | **0.890(0.872-0.906)** | 0.885(0.867-0.903) | **0.572(0.529-0.617)** | 0.553(0.509-0.601) | 0.846(0.816-0.875) | **0.875(0.846-0.904)** | **0.745(0.737-0.754)** | 0.677(0.668-0.687) | p=0.164 |
| UHB | 0.855(0.837-0.873) | **0.858(0.841-0.876)** | **0.353(0.318-0.399)** | 0.315(0.285-0.353) | 0.854(0.825-0.882) | **0.877(0.852-0.903)** | **0.679(0.671-0.687)** | 0.641(0.633-0.649) | p=0.014 |
| BH | **0.920(0.897-0.942)** | 0.917(0.890-0.939) | 0.693(0.630-0.760) | **0.701(0.635-0.763)** | 0.917(0.874-0.954) | **0.924(0.884-0.958)** | **0.734(0.712-0.756)** | 0.713(0.689-0.738) | p=0.379 |
| OUH "wave 2" | 0.867(0.858-0.875) | 0.867(0.858-0.875) | **0.615(0.596-0.634)** | 0.584(0.566-0.604) | 0.865(0.851-0.877) | **0.888(0.876-0.899)** | **0.647(0.641-0.652)** | 0.565(0.56-0.571) | p=0.005 |
| OUH "wave 1" | 0.839(0.823-0.853) | **0.845(0.83-0.86)** | **0.124(0.106-0.146)** | 0.098(0.087-0.11) | 0.850(0.827-0.871) | **0.873(0.852-0.894)** | **0.642(0.639-0.645)** | 0.564(0.562-0.567) | p=0.325 |
| Error in 20% cases |  |  |  |  |  |  |  |  |  |
| PUH | 0.879(0.860-0.898) | **0.882(0.863-0.899)** | 0.471(0.427-0.519) | **0.537(0.492-0.584)** | 0.854(0.823-0.883) | **0.878(0.850-0.904)** | **0.723(0.715-0.733)** | 0.681(0.673-0.691) | p=0.491 |
| UHB | 0.829(0.809-0.850) | **0.851(0.833-0.869)** | 0.272(0.243-0.305) | **0.361(0.322-0.408)** | 0.834(0.803-0.863) | **0.886(0.862-0.910)** | **0.646(0.638-0.654)** | 0.608(0.599-0.616) | p=0.079 |
| BH | 0.925(0.904-0.943) | **0.930(0.908-0.949)** | 0.679(0.614-0.74) | **0.718(0.65-0.792)** | 0.910(0.868-0.947) | **0.924(0.885-0.959)** | **0.718(0.695-0.741)** | 0.702(0.677-0.723) | p=0.01 |
| OUH "wave 2" | 0.855(0.845-0.864) | **0.869(0.86-0.877)** | 0.524(0.504-0.542) | **0.618(0.599-0.638)** | 0.860(0.847-0.873) | **0.892(0.881-0.903)** | **0.606(0.601-0.612)** | 0.548(0.542-0.554) | p=0.303 |
| OUH "wave 1" | 0.819(0.802-0.836) | **0.84(0.825-0.855)** | 0.065(0.058-0.072) | **0.126(0.110-0.147)** | 0.840(0.818-0.863) | **0.876(0.854-0.897)** | **0.607(0.605-0.610)** | 0.571(0.568-0.573) | p=0.038 |
| Error in 30% cases |  |  |  |  |  |  |  |  |  |
| PUH | 0.856(0.834-0.876) | **0.886(0.868-0.902)** | 0.493(0.448-0.544) | **0.516(0.471-0.565)** | 0.865(0.834-0.891) | **0.888(0.861-0.914)** | 0.593(0.583-0.602) | **0.644(0.635-0.653)** | p=0.051 |
| UHB | 0.830(0.809-0.851) | **0.849(0.830-0.868)** | 0.315(0.281-0.356) | **0.371(0.331-0.416)** | **0.868(0.84-0.892)** | 0.863(0.836-0.89) | 0.527(0.519-0.536) | **0.612(0.604-0.621)** | p=0.005 |
| BH | 0.916(0.891-0.937) | **0.920(0.898-0.941)** | 0.675(0.605-0.74) | **0.678(0.609-0.75)** | **0.958(0.928-0.985)** | 0.917(0.874-0.952) | 0.555(0.530-0.581) | **0.696(0.673-0.721)** | p=0.464 |
| OUH "wave 2" | 0.857(0.848-0.866) | **0.867(0.858-0.875)** | **0.581(0.560-0.600)** | 0.577(0.557-0.597) | 0.891(0.879-0.902) | **0.899(0.888-0.910)** | 0.514(0.509-0.520) | **0.520(0.514-0.525)** | p=0.001 |
| OUH "wave 1" | 0.834(0.818-0.849) | **0.835(0.819-0.851)** | **0.121(0.104-0.142)** | 0.116(0.099-0.136) | **0.880(0.858-0.899)** | 0.873(0.852-0.895) | 0.511(0.508-0.514) | **0.524(0.522-0.527)** | p=0.023 |
| Error in 40% cases |  |  |  |  |  |  |  |  |  |
| PUH | 0.856(0.836-0.876) | **0.884(0.865-0.901)** | 0.488(0.443-0.543) | **0.553(0.506-0.598)** | 0.870(0.841-0.898) | **0.870(0.840-0.898)** | 0.548(0.539-0.558) | **0.642(0.634-0.652)** | p=0.014 |
| UHB | 0.823(0.802-0.844) | **0.846(0.826-0.865)** | 0.352(0.313-0.399) | **0.331(0.295-0.375)** | 0.852(0.823-0.879) | **0.875(0.848-0.902)** | 0.576(0.568-0.585) | **0.598(0.589-0.606)** | p=0.027 |
| BH | 0.894(0.863-0.922) | **0.929(0.904-0.949)** | 0.701(0.640-0.760) | **0.7(0.631-0.776)** | 0.903(0.859-0.941) | **0.938(0.903-0.969)** | 0.634(0.612-0.660) | **0.668(0.644-0.693)** | p=0.262 |
| OUH "wave 2" | 0.830(0.819-0.84) | **0.863(0.854-0.871)** | 0.553(0.532-0.572) | **0.577(0.557-0.598)** | 0.852(0.839-0.865) | **0.898(0.886-0.908)** | **0.514(0.508-0.519)** | 0.511(0.505-0.516) | p<0.001 |
| OUH "wave 1" | 0.805(0.788-0.821) | **0.842(0.828-0.857)** | 0.122(0.105-0.144) | **0.09(0.078-0.104)** | 0.84(0.818-0.863) | **0.89(0.871-0.911)** | 0.497(0.494-0.500) | **0.534(0.531-0.537)** | p=0.002 |

As per expectations, the improvement in AUROC is particularly pronounced in the presence of higher levels of label noise within the training set (p-values were generally significant, i.e. *<* 0.05, at most noise ratios above 10%), providing credibility to the fact that NCR indeed plays a crucial role in constraining the model to learn meaningful patterns, rather than simply memorizing noisy data labels. Detailed numerical results for AUROC, AUPRC, sensitivity, and specificity are shown in Table 1 (results for positive predictive value (PPV) and negative predictive value (NPV) can be found in Supplementary Table 7).

In addition to the considered datasets so far, we present results from two additional case studies conducted using the eICU Collaborative Research Database and the Adult (Census Income) Dataset. Again, NCR proved to be effective in addressing label noise at various noise rates thereby showing its utility in healthcare as well as other tabular data domains. Comprehensive results and analysis for these two tasks can be found in Section H of the Supplementary Material.

#### 4.2.2 Analysis of feature embeddings

By utilizing datasets that contain known noise, we have the opportunity to compare the feature similarity among training examples, considering whether they are correctly or incorrectly labeled as belonging to the same or different classes. In an ideal scenario, the distributions of within-class and between-class similarities for clean examples would not overlap and would perfectly match the true within-class and between-class similarities for mislabeled examples. We perform this comparison for both the *baseline NN* model and the model when trained with NCR. Although the distributions for the baseline model exhibit overlap, they are not identical, indicating that the feature similarities still contain some useful information that NCR can leverage. The utilization of NCR during training results in improved separation of classes in the feature space (Figure 3).

We further compare the distribution of cosine similarities for training examples that are correctly or incorrectly labeled as the same class or different classes in Figure 3. The features learned using NCR exhibit noticeably better class separation when compared to the features learned using the *baseline NN* utilizing only cross entropy (CE) loss (top row).

#### 4.2.3 Analysis of prediction confidence

In Figure 4, additional evidence is provided to support the hypothesis that NCR effectively prevents the memorization of noisy labels. Namely, we assessed the confidence level associated with the predicted label for each training example. Panel b) of the figure demonstrates that the model trained with NCR more frequently assigns higher confidence to correctly labeled samples. In contrast, the model trained without NCR, in panel a), tends to overfit to the noisy labels, resulting in a lower confidence assignment to a larger number of correctly labeled samples.

## 5 Conclusion and Discussion

The results of this research highlight the significant promise of incorporating CV methods to tackle the issues stemming from noisy labels in EHR data. Our investigation underscores that despite the notable differences between CV and EHR datasets, the adaptation and fusion of various CV techniques to address label inaccuracies can offer substantial benefits in healthcare applications. Through the utilization of these recently introduced CV techniques, specifically Label Smoothing, Mix-up, and NCR, we can significantly improve the robustness of DL models when confronted with noisy labels during the analysis of EHR data.

Similar to previous studies (38; 42; 55), we noticed that deep neural networks, when trained without these noise-mitigation methods, have the capability to memorize random and noisy labels and exhibit poor generalization on unseen test sets (see Supplementary Table 6 and Figure 1). In particular, we showed that both Mix-up and NCR, individually as well as when combined, demonstrated strong performance in mitigating overfitting to noisy class labels. This success can be attributed to the fact that neither of these methods solely relies on the initial labels (which can be noisy) for evaluating the model’s performance.

We also found that the use of NCR achieves performance that is comparable, and in some cases even superior, to that of standard cross entropy when there is 0% added noise, suggesting a general regularization effect of NCR. This may be because the original training data from the hospitals is already noisy to begin with (even without addition of synthetic noise to the labels), and thus, NCR is able to show improved performance when trained with inherently noisy labels. Improvement in AUROC was generally found to be significant, i.e. *p* < 0.05, across the majority of test sets for noise ratios above 10%. This is reflective of how NCR has greater effect when there is higher levels of label corruption, and less significant effect at low levels of label corruption.

In addition to examining each method in isolation, our findings revealed that combining multiple methods, specifically Mix-up and NCR, exhibited superior performance, outperforming each method when used independently. This improvement stems from the joint effect of Mix-up and NCR in mitigating label noise, with Mix-up also contributing to data augmentation during the training process. This paves the way for future research possibilities, further encouraging the use of multiple CV-based techniques for effectively addressing label noise.

Of the three CV techniques considered, label smoothing performed the worst. We hypothesize that this is because it adds noise to both noisy and clean labels. Thus, as the majority of labels (particularly, COVID-19 negative controls) in our training set are correct, label smoothing can be less effective and actually decrease the accuracy of the model in many cases. Furthermore, previous studies have found that the advantage of label smoothing vanishes when label noise is high (44), which we also observed when the error in COVID-19 positive cases was high.

Our ablation studies with NCR reveal that increasing the weight of NCR can improve performance at higher noise ratios, up to a certain extent. This finding aligns with the expectation that higher noise ratios require a stronger NCR effect to counteract the impact of noise. Additionally, we found that providing a *warm-up* period of few training epochs with just the binary cross-entropy loss enabled (before switching on the NCR loss term) helps achieve superior performance compared to enabling both loss terms from the start of training. We hypothesize that this is because the initial training phase with just the cross entropy loss allows the model to learn the underlying patterns in the noisy data before applying the regularizer, which appears to be more effective when the model has converged to some extent. Also we can view it from a *curriculum learning* perspective, where the model progressively tackles more challenging aspects of the task, i.e., to account for the noise in the data and learning to avoid overfitting to it.

While our efforts were directed at mitigating label noise, it is crucial to emphasize that errors can persist within the features themselves, potentially leading to inaccuracies in the model performance. Feature noise has been a subject of extensive research (45; 32; 34; 9; 13; 27); however, it was not the primary focus of our study. In addition to feature noise, we also encountered missing data concerning the features. To handle this issue, we opted for population median imputation, consistent with similar COVID-19 studies that utilized the same dataset. Nevertheless, it is worth noting that the underlying reasons for and the nature of missing data could carry important information about the source and nature of errors. Therefore, future research should continue exploring alternative approaches for assessing and address missing data.

Moreover, probabilities can prove valuable for specific tasks, particularly when binary classification lacks the necessary depth of information. This approach aligns well with the CV techniques we explored, namely Mix-up and Label Smoothing, both of which yield continuous values instead of discrete binary outcomes. Our decision to employ binary classification (COVID-19 positive or negative) was made to align with rapid triaging processes used in hospitals (36; 53).

Finally, we are also aware that the COVID-19 datasets we employed provide a limited perspective compared to the extensive information available in EHR systems. It is worth noting that significant portions of EHR data, such as treatment-related specifics or lifestyle and environmental factors, among others, are not fully represented in the datasets we have utilized in this study. Therefore, further research is required to gain a comprehensive understanding of the consequences of diverse types of label noise and to assess how different noise mitigation techniques influence model performance.

## Contributions

JY conceived and ran the experiments, with design input from MP. JY and AAS preprocessed the COVID-19 datasets. JY and MP implemented the code. AAS applied for the ethical approval and co-ordinated data extraction. All authors revised the manuscript.

## Acknowledgements

We express our sincere thanks to all patients and staff across the four participating NHS trusts; Oxford University Hospitals NHS Foundation Trust, University Hospitals Birmingham NHS Trust, Bedfordshire Hospitals NHS Foundations Trust, and Portsmouth Hospitals University NHS Trust.

We additionally express our thanks to Artur Speiser and Luca Bertinetto for their comments during this project.

## Ethics

United Kingdom National Health Service (NHS) approval via the national oversight/regulatory body, the Health Research Authority (HRA), has been granted for use of routinely collected clinical data to develop and validate artificial intelligence models to detect Covid-19 (CURIAL; NHS HRA IRAS ID: 281832).

## Data Availability

Data from OUH studied here are available from the Infections in Oxford-shire Research Database (https://oxfordbrc.nihr.ac.uk/research-themes/modernising-medical-microbiology-and-big-infection-diagnostics/infections-in-oxfordshire-research-database-iord/), subject to an application meeting the ethical and governance requirements of the Database. Data from UHB, PUH and BH are available on reasonable request to the respective trusts, subject to HRA requirements.

## Code Availability

Code for this will be available upon publication.

## References

[1] Afzal, A. (2020). Molecular diagnostic technologies for COVID-19: Limitations and challenges. Journal of advanced research, 26, 149–159.

[2] Bell, S. K., Delbanco, T., Elmore, J. G., Fitzgerald, P. S., Fossa, A., Harcourt, K., … & DesRoches, C. M. (2020). Frequency and types of patient-reported errors in electronic health record ambulatory care notes. JAMA network open, 3(6), e205867–e205867.

[3] Ben-Gal, I. (2005). Outlier detection. Data mining and knowledge discovery handbook, 131–146.

[4] Bowman, S. (2013). Impact of electronic health record systems on information integrity: quality and safety implications. Perspectives in health information management, 10(Fall).

[5] Blum, A., & Mitchell, T. (1998, July). Combining labeled and unlabeled data with co-training. In Proceedings of the eleventh annual conference on Computational learning theory (pp. 92–100).

[6] Breunig, M. M., Kriegel, H. P., Ng, R. T., & Sander, J. (2000, May). LOF: identifying densitybased local outliers. In Proceedings of the 2000 ACM SIGMOD international conference on Management of data (pp. 93–104).

[7] Chapman, A. D. (2005). Principles and methods of data cleaning. GBIF.

[8] Chen, T., & Guestrin, C. (2016, August). Xgboost: A scalable tree boosting system. In Proceedings of the 22nd acm sigkdd international conference on knowledge discovery and data mining (pp. 785–794).

[9] Dong, Y., & Peng, C. Y. J. (2013). Principled missing data methods for researchers. Springer-Plus, 2, 1–17.

[10] Ghosh, A., Kumar, H., & Sastry, P. S. (2017, February). Robust loss functions under label noise for deep neural networks. In Proceedings of the AAAI conference on artificial intelligence (Vol. 31, No. 1).

[11] Graber, M. L., Siegal, D., Riah, H., Johnston, D., & Kenyon, K. (2019). Electronic health record–related events in medical malpractice claims. Journal of patient safety, 15(2), 77.

[12] Han, J., Luo, P., & Wang, X. (2019). Deep self-learning from noisy labels. In Proceedings of the IEEE/CVF international conference on computer vision (pp. 5138–5147).

[13] Harel, O., Mitchell, E. M., Perkins, N. J., Cole, S. R., Tchetgen Tchetgen, E. J., Sun, B., & Schisterman, E. F. (2018). Multiple imputation for incomplete data in epidemiologic studies. American journal of epidemiology, 187(3), 576–584.

[14] Iscen, A., Valmadre, J., Arnab, A., & Schmid, C. (2022). Learning with neighbor consistency for noisy labels. In Proceedings of the IEEE/CVF Conference on Computer Vision and Pattern Recognition (pp. 4672–4681).

[15] Kaboli, P. J., McClimon, B. J., Hoth, A. B., & Barnett, M. J. (2004). Assessing the accuracy of computerized medication histories. The American journal of managed care, 10(11 Pt 2), 872–877.

[16] Khajouei, R., Abbasi, R., & Mirzaee, M. (2018). Errors and causes of communication failures from hospital information systems to electronic health record: a record-review study. International journal of medical informatics, 119, 47–53.

[17] Kharrazi, H., Ma, X., Chang, H. Y., Richards, T. M., & Jung, C. (2021). Comparing the predictive effects of patient medication adherence indices in electronic health record and claims-based risk stratification models. Population health management, 24(5), 601–609.

[18] Kim, M. O., Coiera, E., & Magrabi, F. (2017). Problems with health information technology and their effects on care delivery and patient outcomes: a systematic review. Journal of the American Medical Informatics Association, 24(2), 246–250.

[19] Kriegel, H. P., Kröger, P., Schubert, E., & Zimek, A. (2009). Outlier detection in axis-parallel subspaces of high dimensional data. In Advances in Knowledge Discovery and Data Mining: 13th Pacific-Asia Conference, PAKDD 2009 Bangkok, Thailand, April 27-30, 2009 Proceedings 13 (pp. 831–838). Springer Berlin Heidelberg.

[20] Lee, D. H. (2013, June). Pseudo-label: The simple and efficient semi-supervised learning method for deep neural networks. In Workshop on challenges in representation learning, ICML (Vol. 3, No. 2, p. 896).

[21] Ling, Y., An, Y., Liu, M., & Hu, X. (2013, December). An error detecting and tagging framework for reducing data entry errors in electronic medical records (EMR) system. In 2013 IEEE International Conference on Bioinformatics and Biomedicine (pp. 249–254). IEEE.

[22] Lukasik, M., Bhojanapalli, S., Menon, A., & Kumar, S. (2020, November). Does label smoothing mitigate label noise?. In International Conference on Machine Learning (pp. 6448–6458). PMLR.

[23] Menon, S., Singh, H., Giardina, T. D., Rayburn, W. L., Davis, B. P., Russo, E. M., & Sittig, D. F. (2017). Safety huddles to proactively identify and address electronic health record safety. Journal of the American Medical Informatics Association, 24(2), 261–267.

[24] Miller, T. E., Beltran, W. F. G., Bard, A. Z., Gogakos, T., Anahtar, M. N., Astudillo, M. G., … & Lennerz, J. K. (2020). Clinical sensitivity and interpretation of PCR and serological COVID-19 diagnostics for patients presenting to the hospital. The FASEB Journal, 34(10), 13877.

[25] Nigam, K., & Ghani, R. (2000, November). Analyzing the effectiveness and applicability of co-training. In Proceedings of the ninth international conference on Information and knowledge management (pp. 86–93).

[26] Perez, H., & Tah, J. H. (2020). Improving the accuracy of convolutional neural networks by identifying and removing outlier images in datasets using t-SNE. Mathematics, 8(5), 662.

[27] Peskoe, S. B., Arterburn, D., Coleman, K. J., Herrinton, L. J., Daniels, M. J., & Haneuse, S. (2021). Adjusting for selection bias due to missing data in electronic health records-based research. Statistical Methods in Medical Research, 30(10), 2221–2238.

[28] Reed, S., Lee, H., Anguelov, D., Szegedy, C., Erhan, D., & Rabinovich, A. (2014). Training deep neural networks on noisy labels with bootstrapping. arXiv preprint arXiv:1412.6596.

[29] Roman, L. C., Ancker, J. S., Johnson, S. B., & Senathirajah, Y. (2017). Navigation in the electronic health record: a review of the safety and usability literature. Journal of biomedical informatics, 67, 69–79.

[30] Ronquillo, J. G., & Zuckerman, D. M. (2017). Software-related recalls of health information technology and other medical devices: Implications for FDA regulation of digital health. The Milbank Quarterly, 95(3), 535–553.

[31] Saah, A. J., & Hoover, D. R. (1997). “Sensitivity” and “specificity” reconsidered: the meaning of these terms in analytical and diagnostic settings.

[32] Seaman, S. R., & White, I. R. (2013). Review of inverse probability weighting for dealing with missing data. Statistical methods in medical research, 22(3), 278–295.

[33] Staroselsky, M., Volk, L. A., Tsurikova, R., Newmark, L. P., Lippincott, M., Litvak, I., … & Bates, D. W. (2008). An effort to improve electronic health record medication list accuracy between visits: patients’ and physicians’ response. International journal of medical informatics, 77(3), 153–160.

[34] Sterne, J. A., White, I. R., Carlin, J. B., Spratt, M., Royston, P., Kenward, M. G., … & Carpenter, J. R. (2009). Multiple imputation for missing data in epidemiological and clinical research: potential and pitfalls. Bmj, 338.

[35] Stolarek, I., Samelak-Czajka, A., Figlerowicz, M., & Jackowiak, P. (2022). Dimensionality reduction by UMAP for visualizing and aiding in classification of imaging flow cytometry data. Iscience, 25(10).

[36] Soltan, A. A., Yang, J., Pattanshetty, R., Novak, A., Yang, Y., Rohanian, O., … & Muthusami, V. (2022). Real-world evaluation of rapid and laboratory-free COVID-19 triage for emergency care: external validation and pilot deployment of artificial intelligence driven screening. The Lancet Digital Health, 4(4), e266–e278.

[37] Soltan, A. A., Kouchaki, S., Zhu, T., Kiyasseh, D., Taylor, T., Hussain, Z. B., … & Clifton, D. A. (2021). Rapid triage for COVID-19 using routine clinical data for patients attending hospital: development and prospective validation of an artificial intelligence screening test. The Lancet Digital Health, 3(2), e78–e87.

[38] Song, H., Kim, M., Park, D., Shin, Y., & Lee, J. G. (2022). Learning from noisy labels with deep neural networks: A survey. IEEE Transactions on Neural Networks and Learning Systems.

[39] Szegedy, C., Vanhoucke, V., Ioffe, S., Shlens, J., & Wojna, Z. (2016). Rethinking the inception architecture for computer vision. In Proceedings of the IEEE conference on computer vision and pattern recognition (pp. 2818–2826).

[40] Van Stralen, K. J., Stel, V. S., Reitsma, J. B., Dekker, F. W., Zoccali, C., & Jager, K. J. (2009). Diagnostic methods I: sensitivity, specificity, and other measures of accuracy. Kidney international, 75(12), 1257–1263.

[41] Wagner, M. M., & Hogan, W. R. (1996). The accuracy of medication data in an outpatient electronic medical record. Journal of the American Medical Informatics Association, 3(3), 234–244.

[42] Wang, Y., Ma, X., Chen, Z., Luo, Y., Yi, J., & Bailey, J. (2019). Symmetric cross entropy for robust learning with noisy labels. In Proceedings of the IEEE/CVF international conference on computer vision (pp. 322–330).

[43] Wang, X., Hua, Y., Kodirov, E., Clifton, D. A., & Robertson, N. M. (2021). Proselflc: Progressive self label correction for training robust deep neural networks. In Proceedings of the IEEE/CVF conference on computer vision and pattern recognition (pp. 752–761).

[44] Wei, J., Liu, H., Liu, T., Niu, G., Sugiyama, M., & Liu, Y. (2022, June). To Smooth or Not? When Label Smoothing Meets Noisy Labels. In International Conference on Machine Learning (pp. 23589–23614). PMLR.

[45] Wells, B. J., Chagin, K. M., Nowacki, A. S., & Kattan, M. W. (2013). Strategies for handling missing data in electronic health record derived data. Egems, 1(3).

[46] Williams, T. C., Wastnedge, E., McAllister, G., Bhatia, R., Cuschieri, K., Kefala, K., … & Templeton, K. E. (2020). Sensitivity of RT-PCR testing of upper respiratory tract samples for SARS-CoV-2 in hospitalised patients: a retrospective cohort study. Wellcome open research, 5.

[47] Winden, T. J., Chen, E. S., Monsen, K. A., Wang, Y., & Melton, G. B. (2018). Evaluation of flowsheet documentation in the electronic health record for residence, living situation, and living conditions. AMIA Summits on Translational Science Proceedings, 2018, 236.

[48] Xu, Y., Cao, P., Kong, Y., & Wang, Y. (2019). L_dmi: A novel information-theoretic loss function for training deep nets robust to label noise. Advances in neural information processing systems, 32.

[49] Yadav, S., Kazanji, N. KC N., Paudel, S., Falatko, J., Shoichet, S., … & Barnes, M. A. (2017). Comparison of accuracy of physical examination findings in initial progress notes between paper charts and a newly implemented electronic health record. Journal of the American Medical Informatics Association, 24(1), 140–144.

[50] Yang, J., Soltan, A. A., & Clifton, D. A. (2022). Machine learning generalizability across healthcare settings: insights from multi-site COVID-19 screening. npj Digital Medicine, 5(1), 69.

[51] Yang, J., Soltan, A. A., Eyre, D. W., Yang, Y., & Clifton, D. A. (2023). An adversarial training framework for mitigating algorithmic biases in clinical machine learning. npj Digital Medicine, 6(1), 55.

[52] Yang, J., El-Bouri, R., O’Donoghue, O., Lachapelle, A. S., Soltan, A. A., & Clifton, D. A. (2022). Deep Reinforcement Learning for Multi-class Imbalanced Training. arXiv preprint arXiv:2205.12070.

[53] Yang, J., Soltan, A. A., Eyre, D. W., & Clifton, D. A. (2023). Algorithmic fairness and bias mitigation for clinical machine learning with deep reinforcement learning. Nature Machine Intelligence, 1–11.

[54] Zhang, H., Cisse, M., Dauphin, Y. N., & Lopez-Paz, D. (2018, February). mixup: Beyond Empirical Risk Minimization. In International Conference on Learning Representations.

[55] Zhang, C., Bengio, S., Hardt, M., Recht, B., & Vinyals, O. (2021). Understanding deep learning (still) requires rethinking generalization. Communications of the ACM, 64(3), 107–115.

[56] Boughorbel, S., Fethi J., Neethu V., & Haithum E. (2018). Alternating loss correction for preterm-birth prediction from ehr data with noisy labels. arXiv preprint arXiv:1811.09782 (2018).

[57] Tjandra, D., & Wiens J. Leveraging an Alignment Set in Tackling Instance-Dependent Label Noise. Conference on Health, Inference, and Learning, (2023), 477–497.

[58] Deng J, Dong W, Socher R, Li LJ, Li K, Fei-Fei L. Imagenet: A large-scale hierarchical image database. In2009 IEEE conference on computer vision and pattern recognition 2009 Jun 20 (pp. 248–255). Ieee.

