## Supplementary Material for "Addressing Label Noise for Electronic Health Records: Insights from Computer Vision for Tabular Data"

#### A Software Packages and Implementation

Models implemented using Python (v3.6.9). Scikit Learn (v0.24.1) was used for standardization, imputation, and calculating performance metrics. Neural networks were implemented using PyTorch (v1.13.1). Models run using an Intel Xeon E-2146G Processor (CPU: 6 cores, 4.50 GHz max frequency).

#### B Model Architectures

**Neural Network Model**: The rectified linear unit (ReLU) activation function was used for the hidden layers and the softmax activation function was used in the output layer. For updating model weights, the Adaptive Moment Estimation (Adam) optimizer was used during training.

```
CovidClassifier(
  (hidden1): Linear(in_features=26, out_features=10, bias=True)
  (act1): ReLU()
  (hidden2): Linear(in_features=10, out_features=10, bias=True)
  (act2): ReLU()
  (hidden3): Linear(in_features=10, out_features=10, bias=True)
  (act3): ReLU()
  (hidden4): Linear(in_features=10, out_features=10, bias=True)
  (act4): ReLU()
  (output): Linear(in_features=10, out_features=2, bias=True)
  (act_output): Softmax(dim=None)
)
```

Supplementary Figure 1: Final neural network architecture used

#### C COVID-19 Data and Preprocessing

The following inclusions and exclusions are reproduced from previous studies (Soltan et al., 2022, Yang et al., 2022a, Yang et al., 2022b).

**Oxford University Hospitals NHS Foundation Trust (OUH):** We included all patients attending acute and emergency care settings at OUH who received routine blood tests on arrival, considering presentations before December 1, 2019, and thus before the pandemic, as the COVID-19-negative (control) cohort. We considered presentations during the 'first wave' of the UK COVID-19 pandemic (December 1, 2019 to June 30, 2020) with PCR confirmed SARS-CoV-2 infection as the COVID-19-positive (cases) cohort. We excluded patients who opted out of electronic health record (EHR) research and those who did not receive laboratory blood tests or were younger than 18 years of age. Due to incomplete penetrance of testing during the first wave of the pandemic, and imperfect sensitivity of the PCR test, there is uncertainty in the viral status of patients presenting during the pandemic who were untested or tested negative. We therefore selected a pre-pandemic control cohort during training to ensure absence of disease in patients labelled as COVID-19-negative. Clinical features extracted for each presentation included first-performed blood tests, blood gases, vital signs measurements and PCR testing for SARS-CoV-2 (Abbott Architect [Abbott, Maidenhead, UK], TaqPath [Thermo Fisher Scientific, Massachusetts, USA] and Public Health England-designed RNA-dependent RNA polymerase assays).

**Portsmouth Hospitals University NHS Foundation Trust (PUH):** PUH considered all patients admitted to the Queen Alexandria Hospital, serving a population of 675,000 and offering tertiary referral services to the surrounding region, between March 1, 2020 and February 28, 2021. Confirmatory COVID-19 testing was by laboratory SARS-CoV2 RT-PCR assay, considering any positive PCR result within 48hrs of admission as a true positive.

**University Hospitals Birmingham NHS Foundation Trust (UHB):** UHB considered all patients admitted to The Queen Elizabeth Hospital, Birmingham, between December 01, 2019 and October 29, 2020. The Queen Elizabeth Hospital is a large tertiary referral unit within the UHB group which provides healthcare services for a population of 2.2 million across the West Midlands. Confirmatory COVID-19 testing was performed by laboratory SARS-CoV-2 RT-PCR assay.

**Bedfordshire NHS Foundation Trust (BH):** BH considered all patients admitted to Bedford Hospital between January 1, 2021 and March 31, 2021. BH provides healthcare services for a population of around 620,000 in Bedfordshire. Confirmatory COVID-19 testing was performed on the day of admission by point-of-care PCR based nucleic acid testing [SAMBA-II & Panther Fusion System, Diagnostics in the Real World, UK, and Hologic, USA].

Supplementary Table 1: Clinical predictors considered for COVID-19 status prediction.

| **Category** | **Features** |
|---|---|
| Vital Signs | Heart rate, respiratory rate, oxygen saturation, systolic blood pressure, diastolic blood pressure, temperature |
| Blood Tests | Haemoglobin, haematocrit, mean cell volume, white cell count, neutrophil count, lymphocyte count, monocyte count, eosinophil count, basophil count, platelets |
| Liver Function Tests & C-reactive protein | Albumin, alkaline phosphatase, alanine aminotransferase, bilirubin, C-reactive protein |
| Urea & Electrolytes | Sodium, potassium, creatinine, urea, estimated glomerular filtration rate |

Supplementary Table 2: Summary of number of patients, COVID-19 positive cases

| | Training | Validation | Test |
|---|---|---|---|
| Total patients | 22,737 (1,182 positive) | 7,579 (439 positive) | 148,470 (4,226 positive) |
| PUH | 22,737 (1,182 positive) | 7,579 (439 positive) | 7,580 (384 positive) |
| UHB | NA | NA | 10,293 (439 positive) |
| BH | NA | NA | 1,177 (144 positive) |
| OUH "wave 2" | NA | NA | 22,857 (2,012 positive) |
| OUH "wave 1" | NA | NA | 92,671 (701 positive) |

#### D Comparison of Methods

Supplementary Table 3: Hyperparameter values for final models presented in main text.

| | 0% Error | 10% Error in Cases, 0.5% Error in Controls | 20% Error in Cases, 0.5% Error in Controls | 30% Error in Cases, 0.5% Error in Controls | 40% Error in Cases, 0.5% Error in Controls |
|---|---|---|---|---|---|
| CE | | | | | |
| Epochs | 100 | 100 | 100 | 100 | 100 |
| Batch | 2048 | 2048 | 2048 | 2048 | 2048 |
| XGBoost | | | | | |
| Depth | 3 | 3 | 3 | 3 | 3 |
| Label Smoothing | | | | | |
| Epsilon | 0.1 | 0.1 | 0.2 | 0.1 | 0.2 |
| Mix-Up | | | | | |
| Probability of mix-up | 0.3 | 0.3 | 0.3 | 0.3 | 0.3 |
| Alpha | 0.2 | 0.5 | 0.5 | 0.4 | 0.5 |
| NCR | | | | | |
| Epochs | 100 | 100 | 100 | 100 | 100 |
| Batch | 2048 | 2048 | 2048 | 2048 | 2048 |
| NCR Starting Epoch | 30 | 30 | 30 | 30 | 30 |
| Hidden Layer (for NCR) | 1 | 1 | 1 | 1 | 1 |
| NCR weight | 0.05 | 0.03 | 0.03 | 0.04 | 0.03 |
| k | 10 | 10 | 10 | 10 | 10 |

Supplementary Table 4: Comparison of mean AUROC performances (alongside standard deviation) for each test set, across different comparators. Red and blue values denote the best and second best performing methods for each test set, respectively.

| Test Set | Baseline NN | | XGBoost | | Label Smoothing | | Mix-up | | Mix-up + NCR | | NCR | |
|---|---|---|---|---|---|---|---|---|---|---|---|---|
| | Mean | Std. | Mean | Std. | Mean | Std. | Mean | Std. | Mean | Std. | Mean | Std. |
| PUH | 0.873 | 0.016 | 0.875 | 0.010 | 0.869 | 0.013 | 0.888 | 0.014 | 0.893 | **0.004** | 0.886 | **0.005** |
| UHB | 0.839 | 0.016 | 0.824 | 0.014 | 0.843 | 0.011 | 0.853 | **0.006** | 0.856 | 0.008 | 0.852 | **0.005** |
| BH | 0.915 | 0.012 | 0.896 | 0.014 | 0.923 | 0.012 | 0.933 | **0.005** | 0.925 | **0.005** | 0.926 | **0.007** |
| OUH "wave 2" | 0.855 | 0.015 | 0.844 | 0.011 | 0.859 | 0.010 | 0.868 | **0.005** | 0.869 | **0.005** | 0.869 | **0.006** |
| OUH "wave 1" | 0.828 | 0.016 | 0.805 | 0.031 | 0.838 | 0.012 | 0.842 | 0.012 | 0.843 | **0.005** | 0.841 | **0.004** |

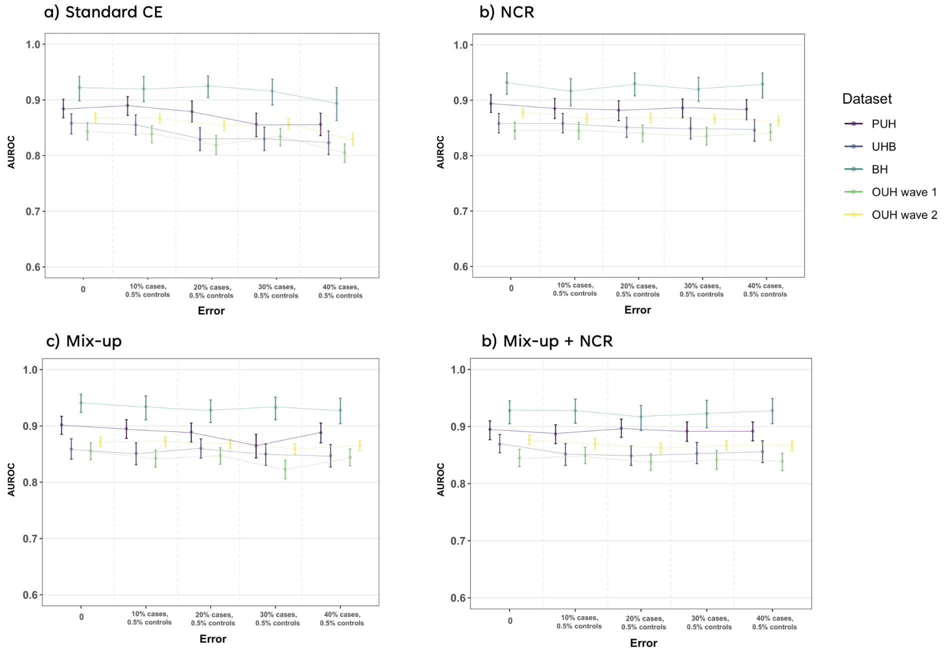


Supplementary Figure 2: Change in AUROC at different training error levels. Panel a) shows results when models are trained using standard cross-entropy, panel b) shows results when models were trained with NCR, panel c) shows results when models were trained with Mix-up, and panel d) shows results when models were trained using a combination of Mix-up and NCR.

Supplementary Table 5: p-values shown compare differences in performance between the Mix-up and Mix-up+NCR methods to the NCR model. p-values are obtained through 1,000 bootstrapped iterations.

| Error in Training Set | Test Set | Mix-up | Mix-up+NCR |
|---|---|---|---|
| 0 | PUH | 0.356 | 0.441 |
| | UHB | 0.007 | 0.056 |
| | BH | 0.095 | 0.314 |
| | OUH2 | 0.131 | 0.072 |
| | OUH1 | 0.032 | 0.378 |
| 10% cases, 0.5% controls | PUH | 0.105 | 0.149 |
| | UHB | 0.072 | 0.278 |
| | BH | 0.208 | 0.27 |
| | OUH2 | <0.001 | 0.102 |
| | OUH1 | 0.156 | 0.086 |
| 20% cases, 0.5% controls | PUH | 0.024 | 0.442 |
| | UHB | 0.021 | 0.36 |
| | BH | 0.105 | 0.177 |
| | OUH2 | 0.27 | 0.346 |
| | OUH1 | 0.032 | 0.396 |
| 30% cases, 0.5% controls | PUH | 0.031 | 0.019 |
| | UHB | 0.181 | 0.1 |
| | BH | 0.026 | 0.453 |
| | OUH2 | 0.043 | 0.088 |
| | OUH1 | 0.475 | 0.317 |
| 40%, cases 0.5% controls | PUH | 0.089 | 0.38 |
| | UHB | 0.101 | 0.083 |
| | BH | 0.045 | 0.073 |
| | OUH2 | 0.001 | 0.175 |
| | OUH1 | 0.045 | 0.052 |

### E Additional Results

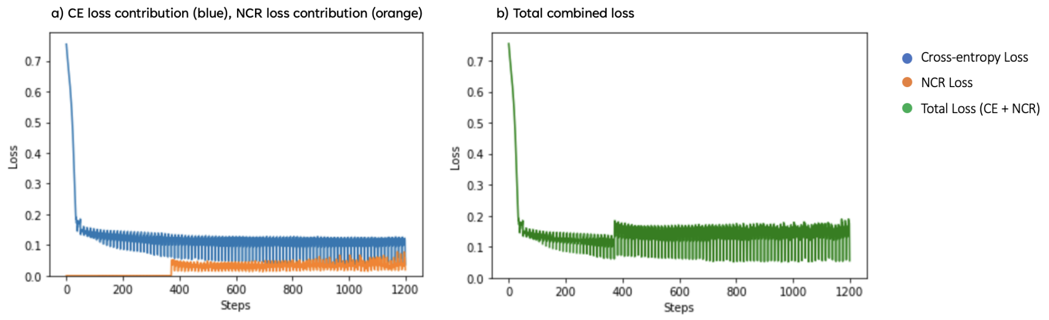


Supplementary Figure 3: Loss during NCR model training. Panel a) shows CE and NCR loss contributions separately (CE:NCR is about 10:3 ratio), and panel b) shows the combined total loss.

Supplementary Table 6: **AUROC comparison of different methods across different amounts of error (i.e. label corruption) for all considered test sets.** CV-based methods are highlighted in bold. In addition to label error in COVID-19 positive cases, there is also 0.5% label error in the negative controls. 0% error represents the original dataset, without any added label noise. Red and blue values denote the best and second best performing methods for each test set, respectively.

| Test Set | Baseline NN | XGBoost | **Label Smoothing** | **Mix-up** | **NCR** | **Mix-up + NCR** |
|---|---|---|---|---|---|---|
| 0% error | | | | | | |
| PUH | 0.884(0.868-0.901) | 0.892(0.873-0.908) | 0.877(0.858-0.895) | **0.902(0.885-0.917)** | 0.894(0.878-0.91) | **0.895(0.877-0.91)** |
| UHB | 0.858(0.839-0.875) | 0.841(0.823-0.859) | 0.854(0.834-0.873) | **0.859(0.841-0.877)** | 0.858(0.841-0.876) | **0.869(0.854-0.886)** |
| BH | 0.922(0.898-0.942) | 0.914(0.89-0.933) | 0.932(0.915-0.949) | **0.941(0.924-0.956)** | **0.932(0.911-0.949)** | 0.928(0.905-0.945) |
| OUH "wave 2" | 0.868(0.86-0.877) | 0.856(0.847-0.865) | 0.87(0.861-0.877) | 0.872(0.863-0.88) | **0.878(0.869-0.885)** | **0.877(0.868-0.885)** |
| OUH "wave 1" | 0.843(0.828-0.859) | **0.856(0.847-0.865)** | 0.848(0.834-0.863) | **0.855(0.84-0.87)** | 0.845(0.83-0.86) | 0.845(0.83-0.86) |
| Error in 10% cases | | | | | | |
| PUH | **0.89(0.872-0.906)** | 0.872(0.852-0.892) | 0.876(0.858-0.894) | **0.895(0.878-0.911)** | 0.885(0.867-0.903) | 0.887(0.87-0.903) |
| UHB | **0.855(0.837-0.873)** | 0.838(0.819-0.854) | 0.843(0.823-0.862) | 0.851(0.831-0.87) | **0.858(0.841-0.876)** | 0.852(0.832-0.87) |
| BH | 0.92(0.897-0.942) | 0.907(0.884-0.929) | 0.924(0.902-0.944) | **0.934(0.911-0.953)** | 0.917(0.89-0.939) | **0.928(0.906-0.948)** |
| OUH "wave 2" | 0.867(0.858-0.875) | 0.85(0.84-0.858) | 0.858(0.848-0.867) | **0.873(0.864-0.881)** | 0.867(0.858-0.875) | **0.87(0.861-0.879)** |
| OUH "wave 1" | 0.839(0.823-0.853) | 0.795(0.778-0.813) | 0.83(0.815-0.846) | 0.842(0.827-0.857) | **0.845(0.83-0.86)** | **0.849(0.835-0.864)** |
| Error in 20% cases | | | | | | |
| PUH | 0.879(0.86-0.898) | 0.875(0.857-0.894) | 0.876(0.857-0.893) | **0.889(0.871-0.905)** | 0.882(0.863-0.899) | **0.897(0.881-0.913)** |
| UHB | 0.829(0.809-0.85) | 0.821(0.803-0.841) | 0.836(0.816-0.855) | **0.86(0.843-0.877)** | **0.851(0.833-0.869)** | 0.849(0.832-0.866) |
| BH | 0.925(0.904-0.943) | 0.895(0.871-0.917) | 0.919(0.896-0.939) | **0.928(0.906-0.946)** | **0.93(0.908-0.949)** | 0.917(0.893-0.937) |
| OUH "wave 2" | 0.855(0.845-0.864) | 0.85(0.84-0.858) | **0.864(0.855-0.873)** | **0.869(0.86-0.877)** | **0.869(0.86-0.877)** | 0.863(0.854-0.871) |
| OUH "wave 1" | 0.819(0.802-0.836) | 0.807(0.791-0.824) | **0.846(0.832-0.861)** | **0.847(0.832-0.861)** | 0.84(0.825-0.855) | 0.838(0.823-0.852) |
| Error in 30% cases | | | | | | |
| PUH | 0.856(0.834-0.876) | 0.867(0.848-0.887) | 0.869(0.851-0.888) | 0.865(0.843-0.885) | **0.886(0.868-0.902)** | **0.892(0.874-0.908)** |
| UHB | 0.83(0.809-0.851) | 0.809(0.789-0.828) | **0.855(0.838-0.872)** | 0.85(0.83-0.868) | 0.849(0.83-0.868) | **0.853(0.835-0.872)** |
| BH | 0.916(0.891-0.937) | 0.883(0.852-0.909) | **0.935(0.916-0.951)** | **0.933(0.911-0.951)** | 0.92(0.898-0.941) | 0.923(0.898-0.946) |
| OUH "wave 2" | 0.857(0.848-0.866) | 0.837(0.827-0.846) | 0.858(0.849-0.867) | **0.86(0.85-0.868)** | **0.867(0.858-0.875)** | **0.867(0.858-0.875)** |
| OUH "wave 1" | 0.834(0.818-0.849) | 0.792(0.776-0.808) | **0.845(0.83-0.86)** | 0.823(0.806-0.839) | 0.835(0.819-0.851) | **0.842(0.825-0.858)** |
| Error in 40% cases | | | | | | |
| PUH | 0.856(0.836-0.876) | 0.871(0.85-0.89) | 0.847(0.823-0.869) | **0.888(0.87-0.905)** | 0.884(0.865-0.901) | **0.892(0.875-0.908)** |
| UHB | 0.823(0.802-0.844) | 0.813(0.793-0.831) | 0.829(0.807-0.849) | **0.847(0.827-0.867)** | 0.846(0.826-0.865) | **0.856(0.837-0.875)** |
| BH | 0.894(0.863-0.922) | 0.882(0.854-0.908) | 0.905(0.88-0.929) | **0.928(0.904-0.949)** | **0.929(0.904-0.949)** | **0.928(0.905-0.949)** |
| OUH "wave 2" | 0.83(0.819-0.84) | 0.828(0.818-0.837) | 0.844(0.835-0.854) | **0.865(0.856-0.874)** | 0.863(0.854-0.871) | **0.867(0.857-0.875)** |
| OUH "wave 1" | 0.805(0.788-0.821) | 0.773(0.756-0.792) | 0.82(0.803-0.837) | **0.844(0.829-0.859)** | **0.842(0.828-0.857)** | 0.839(0.823-0.853) |

Supplementary Table 7: PPV and NPV comparison between baseline and NCR models, across different amounts of error and test sets. In addition to label error in COVID-19 positive cases, there is also 0.5% label error in the negative controls. 0% error represents the original dataset, without any added label noise.

| Test Set | PPV | | NPV | |
|---|---|---|---|---|
| | CE | CE+NCR | CE | CE+NCR |
| 0% error | | | | |
| PUH | 0.139(0.133-0.145) | **0.203(0.193-0.213)** | 0.988(0.986-0.991) | 0.988(0.986-0.990) |
| UHB | 0.096(0.092-0.099) | **0.118(0.113-0.123)** | 0.990(0.988-0.992) | **0.990(0.988-0.991)** |
| BH | 0.276(0.261-0.292) | **0.353(0.33-0.378)** | **0.986(0.978-0.992)** | 0.983(0.975-0.989) |
| OUH "wave 2" | 0.176(0.173-0.179) | **0.224(0.22-0.228)** | 0.980(0.978-0.982) | 0.980(0.978-0.981) |
| OUH "wave 1" | 0.017(0.016-0.017) | **0.022(0.021-0.023)** | 0.998(0.998-0.999) | 0.998(0.998-0.998) |
| Error in 10% cases | | | | |
| PUH | **0.150(0.144-0.157)** | 0.126(0.121-0.131) | 0.989(0.987-0.991) | **0.990(0.988-0.992)** |
| UHB | **0.106(0.102-0.11)** | 0.098(0.095-0.101) | 0.991(0.989-0.992) | **0.992(0.990-0.993)** |
| BH | **0.324(0.305-0.346)** | 0.310(0.290-0.330) | 0.984(0.977-0.991) | **0.985(0.978-0.992)** |
| OUH "wave 2" | **0.191(0.188-0.194)** | 0.165(0.162-0.167) | 0.980(0.978-0.982) | **0.981(0.979-0.983)** |
| OUH "wave 1" | **0.018(0.017-0.018)** | 0.015(0.015-0.015) | 0.998(0.998-0.998) | 0.998(0.998-0.999) |
| Error in 20% cases | | | | |
| PUH | **0.142(0.136-0.147)** | 0.128(0.123-0.133) | 0.989(0.987-0.991) | **0.991(0.988-0.993)** |
| UHB | **0.095(0.091-0.099)** | 0.091(0.088-0.094) | 0.989(0.987-0.991) | **0.992(0.990-0.993)** |
| BH | **0.310(0.291-0.331)** | 0.302(0.284-0.320) | 0.983(0.975-0.990) | **0.985(0.978-0.992)** |
| OUH "wave 2" | **0.174(0.171-0.177)** | 0.160(0.157-0.162) | 0.978(0.976-0.980) | **0.981(0.979-0.983)** |
| OUH "wave 1" | **0.016(0.016-0.016)** | 0.015(0.015-0.016) | 0.998(0.998-0.998) | 0.998(0.998-0.999) |
| Error in 30% cases | | | | |
| PUH | 0.102(0.098-0.105) | **0.117(0.113-0.122)** | 0.988(0.985-0.990) | **0.991(0.989-0.993)** |
| UHB | 0.076(0.073-0.078) | **0.090(0.087-0.093)** | 0.989(0.987-0.991) | **0.990(0.988-0.992)** |
| BH | 0.231(0.219-0.242) | **0.296(0.278-0.314)** | **0.990(0.982-0.996)** | 0.984(0.976-0.991) |
| OUH "wave 2" | 0.150(0.148-0.153) | **0.153(0.151-0.155)** | 0.980(0.978-0.982) | **0.982(0.980-0.983)** |
| OUH "wave 1" | 0.014(0.013-0.014) | 0.014(0.013-0.014) | 0.998(0.998-0.999) | 0.998(0.998-0.998) |
| Error in 40% cases | | | | |
| PUH | 0.093(0.09-0.097) | **0.115(0.111-0.119)** | 0.987(0.985-0.99) | **0.989(0.987-0.992)** |
| UHB | 0.082(0.079-0.085) | **0.088(0.085-0.091)** | 0.989(0.986-0.991) | **0.991(0.989-0.993)** |
| BH | 0.256(0.241-0.274) | **0.282(0.266-0.300)** | 0.979(0.970-0.987) | **0.987(0.980-0.994)** |
| OUH "wave 2" | 0.145(0.142-0.147) | **0.150(0.148-0.153)** | 0.973(0.971-0.975) | **0.981(0.979-0.983)** |
| OUH "wave 1" | 0.013(0.012-0.013) | **0.014(0.014-0.015)** | 0.998(0.997-0.998) | **0.998(0.998-0.999)** |

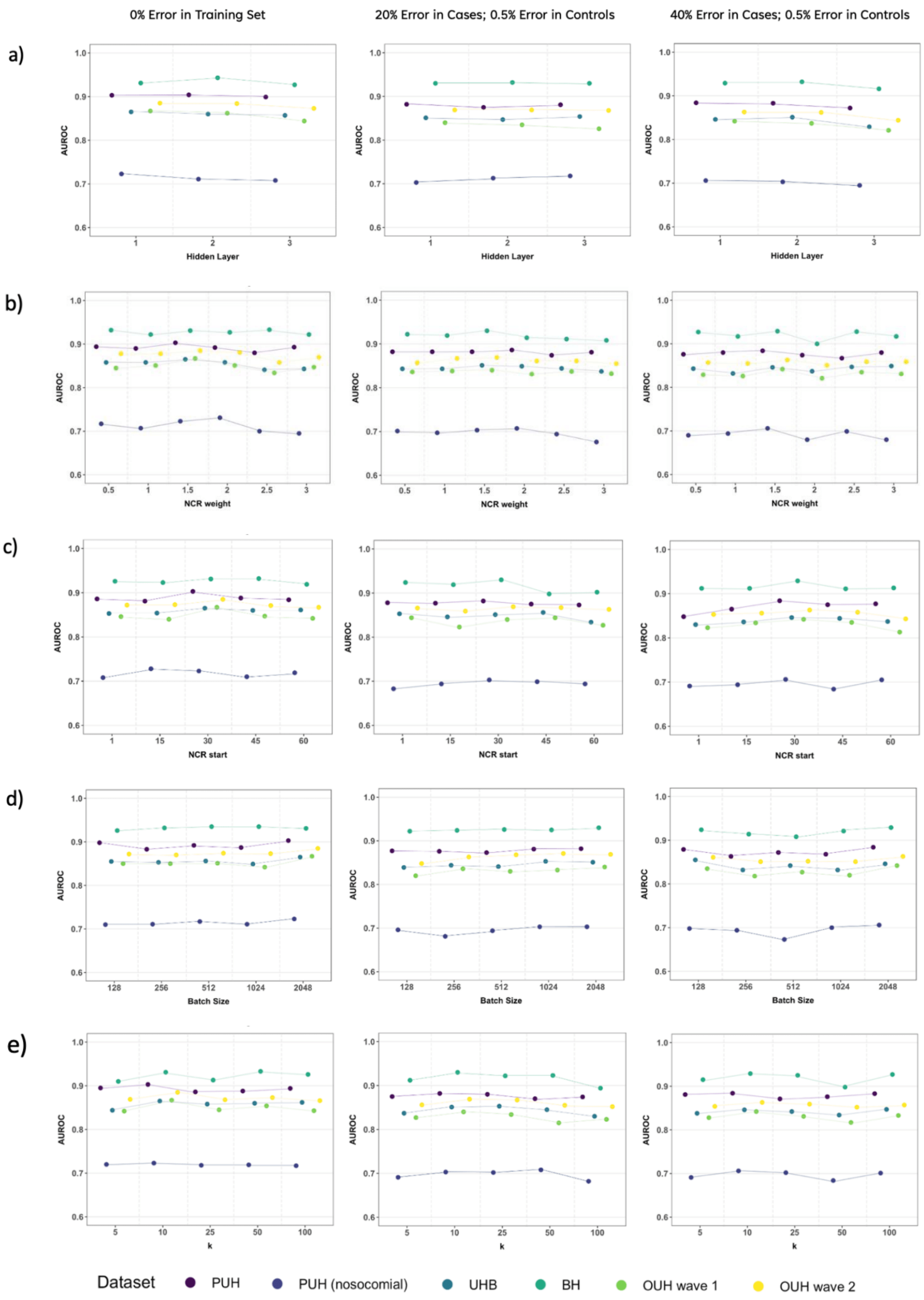


Supplementary Figure 4: Ablation study across varying hidden layer size, NCR weight, NCR starting epoch, batch size, and number of nearest neighbours (k). Results presented for 0% error, 20% error in cases and 0.5% error in controls, and 40% error in cases and 0.5% error in controls.

### F Changing the Loss function

#### F.1 Jensen-Shannon Divergence

$$L_{NCR} := \frac{1}{m}\sum_{i=1}^{m} D_{JS}\left(\sigma(\mathbf{z_i}) \middle\| \sum_{j\epsilon NN_k} \frac{s_{i,j}}{\sum_k s_{i,k}} \sigma(\mathbf{z_j})\right) \tag{1}$$

Supplementary Table 8: AUROC, AUPRC, Sensitivity, and Specificity comparison between baseline and NCR models, across different amounts of error and test sets. In addition to label error in COVID-19 positive cases, there is also 0.5% label error in the negative controls. 0% error represents the original dataset, without any added label noise.

| Test Set | AUROC | | AUPRC | | Sensitivity | | Specificity | |
|---|---|---|---|---|---|---|---|---|
| | CE | CE+NCR | CE | CE+NCR | CE | CE+NCR | CE | CE+NCR |
| 0% error | | | | | | | | |
| PUH | 0.884(0.868-0.901) | **0.885(0.867-0.903)** | 0.538(0.494-0.583) | **0.588(0.546-0.630)** | **0.841(0.810-0.871)** | 0.820(0.789-0.850) | 0.722(0.713-0.731) | **0.810(0.803-0.817)** |
| UHB | 0.858(0.839-0.875) | **0.865(0.846-0.882)** | 0.309(0.278-0.34) | **0.417(0.375-0.461)** | **0.861(0.833-0.889)** | 0.852(0.823-0.880) | 0.638(0.629-0.646) | **0.717(0.710-0.725)** |
| BH | 0.922(0.898-0.942) | **0.935(0.916-0.951)** | 0.691(0.627-0.751) | **0.743(0.682-0.801)** | **0.931(0.894-0.961)** | 0.910(0.872-0.949) | 0.659(0.636-0.684) | **0.758(0.737-0.781)** |
| OUH "wave 2" | 0.868(0.860-0.877) | **0.888(0.872-0.888)** | 0.554(0.536-0.573) | **0.663(0.645-0.680)** | **0.875(0.862-0.887)** | 0.854(0.839-0.867) | 0.604(0.598-0.609) | **0.703(0.698-0.708)** |
| OUH "wave 1" | 0.843(0.828-0.859) | **0.855(0.841-0.870)** | 0.087(0.078-0.097) | **0.180(0.154-0.210)** | **0.859(0.837-0.882)** | 0.810(0.785-0.836) | 0.615(0.613-0.618) | **0.721(0.719-0.724)** |
| Error in 10% cases | | | | | | | | |
| PUH | 0.890(0.872-0.906) | **0.892(0.875-0.909)** | **0.572(0.529-0.617)** | 0.526(0.478-0.580) | 0.846(0.816-0.875) | **0.865(0.835-0.893)** | 0.745(0.737-0.754) | **0.758(0.750-0.766)** |
| UHB | **0.855(0.837-0.873)** | 0.854(0.834-0.872) | 0.353(0.318-0.399) | 0.353(0.319-0.396) | **0.854(0.825-0.882)** | 0.836(0.805-0.865) | 0.679(0.671-0.687) | **0.702(0.694-0.709)** |
| BH | 0.920(0.897-0.942) | **0.926(0.901-0.947)** | 0.693(0.630-0.760) | **0.713(0.649-0.774)** | 0.917(0.874-0.954) | **0.924(0.885-0.959)** | 0.734(0.712-0.756) | **0.771(0.748-0.792)** |
| OUH "wave 2" | 0.867(0.858-0.875) | **0.869(0.861-0.877)** | **0.615(0.596-0.634)** | 0.591(0.571-0.611) | 0.865(0.851-0.877) | **0.866(0.853-0.878)** | 0.647(0.641-0.652) | **0.653(0.647-0.658)** |
| OUH "wave 1" | 0.839(0.823-0.853) | **0.841(0.826-0.856)** | **0.124(0.106-0.146)** | 0.106(0.093-0.123) | **0.850(0.827-0.871)** | 0.845(0.821-0.867) | 0.642(0.639-0.645) | **0.653(0.650-0.656)** |
| Error in 20% cases | | | | | | | | |
| PUH | 0.879(0.860-0.898) | **0.887(0.869-0.902)** | 0.471(0.427-0.519) | **0.495(0.453-0.545)** | 0.854(0.823-0.883) | **0.880(0.852-0.906)** | **0.723(0.715-0.733)** | 0.656(0.647-0.665) |
| UHB | 0.829(0.809-0.850) | **0.839(0.819-0.859)** | 0.272(0.243-0.305) | **0.322(0.289-0.364)** | 0.834(0.803-0.863) | **0.841(0.810-0.869)** | **0.646(0.638-0.654)** | 0.636(0.627-0.644) |
| BH | **0.925(0.904-0.943)** | 0.914(0.888-0.936) | **0.679(0.614-0.74)** | 0.649(0.582-0.725) | 0.910(0.868-0.947) | **0.931(0.891-0.964)** | **0.718(0.695-0.741)** | 0.711(0.688-0.735) |
| OUH "wave 2" | 0.855(0.845-0.864) | **0.867(0.857-0.875)** | 0.524(0.504-0.542) | **0.583(0.563-0.602)** | 0.860(0.847-0.873) | **0.876(0.862-0.887)** | 0.606(0.601-0.612) | **0.609(0.603-0.614)** |
| OUH "wave 1" | 0.819(0.802-0.836) | **0.832(0.815-0.848)** | 0.065(0.058-0.072) | **0.108(0.095-0.123)** | 0.840(0.818-0.863) | **0.846(0.824-0.869)** | **0.607(0.605-0.610)** | 0.594(0.592-0.597) |
| Error in 30% cases | | | | | | | | |
| PUH | 0.856(0.834-0.876) | **0.887(0.87-0.903)** | 0.493(0.448-0.544) | **0.527(0.482-0.578)** | 0.865(0.834-0.891) | **0.885(0.858-0.912)** | 0.593(0.583-0.602) | **0.628(0.619-0.637)** |
| UHB | 0.830(0.809-0.851) | **0.858(0.84-0.875)** | **0.315(0.281-0.356)** | 0.312(0.280-0.353) | 0.868(0.84-0.892) | **0.911(0.889-0.933)** | **0.527(0.519-0.536)** | 0.524(0.515-0.532) |
| BH | 0.916(0.891-0.937) | **0.924(0.898-0.946)** | 0.675(0.605-0.74) | **0.697(0.626-0.770)** | **0.958(0.928-0.985)** | 0.938(0.902-0.969) | 0.555(0.530-0.581) | **0.622(0.598-0.649)** |
| OUH "wave 2" | 0.857(0.848-0.866) | **0.867(0.858-0.876)** | **0.581(0.560-0.60)** | 0.576(0.555-0.598) | 0.891(0.879-0.902) | **0.911(0.900-0.921)** | **0.514(0.509-0.520)** | 0.469(0.463-0.474) |
| OUH "wave 1" | 0.834(0.818-0.849) | **0.848(0.833-0.862)** | **0.121(0.104-0.142)** | 0.082(0.072-0.095) | 0.880(0.858-0.899) | **0.910(0.893-0.928)** | **0.511(0.508-0.514)** | 0.496(0.493-0.499) |
| Error in 40% cases | | | | | | | | |
| PUH | 0.856(0.836-0.876) | **0.883(0.864-0.900)** | 0.488(0.443-0.543) | **0.544(0.501-0.592)** | 0.870(0.841-0.898) | **0.901(0.876-0.925)** | 0.548(0.539-0.558) | **0.577(0.567-0.586)** |
| UHB | 0.823(0.802-0.844) | **0.834(0.813-0.855)** | **0.352(0.313-0.399)** | 0.318(0.283-0.356) | 0.852(0.823-0.879) | **0.877(0.850-0.905)** | **0.576(0.568-0.585)** | 0.546(0.538-0.554) |
| BH | 0.894(0.863-0.922) | **0.921(0.896-0.942)** | 0.701(0.640-0.760) | **0.712(0.640-0.776)** | 0.903(0.859-0.941) | **0.938(0.903-0.969)** | **0.634(0.612-0.660)** | 0.597(0.572-0.623) |
| OUH "wave 2" | 0.830(0.819-0.840) | **0.851(0.841-0.859)** | 0.553(0.532-0.572) | **0.554(0.533-0.573)** | 0.852(0.839-0.865) | **0.901(0.889-0.912)** | **0.514(0.508-0.519)** | 0.460(0.454-0.465) |
| OUH "wave 1" | 0.805(0.788-0.821) | **0.821(0.805-0.837)** | **0.122(0.105-0.144)** | 0.094(0.082-0.107) | 0.840(0.818-0.863) | **0.874(0.853-0.894)** | **0.497(0.494-0.500)** | 0.460(0.457-0.463) |

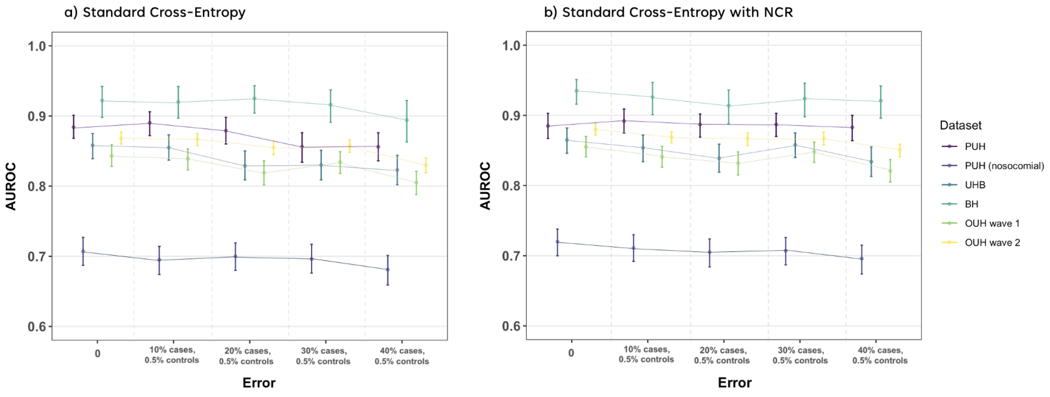


Supplementary Figure 5: Change in performance (AUROC) at different training error levels. Panel a) shows results when models are trained using standard cross-entropy, and panel b) shows results when models were trained with NCR.

Supplementary Table 9: Comparison of mean AUROC performance across different training set error levels.

| Test Set | CE | | CE + NCR | |
|---|---|---|---|---|
| | Mean | Std. | Mean | Std. |
| PUH | 0.873 | 0.016 | **0.887** | **0.003** |
| UHB | 0.839 | 0.016 | **0.850** | **0.013** |
| BH | 0.915 | 0.012 | **0.924** | **0.008** |
| OUH "wave 2" | 0.855 | 0.015 | **0.867** | **0.010** |
| OUH "wave 1" | 0.828 | 0.016 | **0.839** | **0.013** |

Supplementary Table 10: Hyperparameter values for final models (NCR term based on JS divergence) presented in main text.

| Loss Function | CE | CE + NCR | | | | |
|---|---|---|---|---|---|---|
| Error in Training Labels | 0% Error | 0% Error | 10% Error in Cases, 0.5% Error in Controls | 20% Error in Cases, 0.5% Error in Controls | 30% Error in Cases, 0.5% Error in Controls | 40% Error in Cases, 0.5% Error in Controls |
| Epochs | 100 | 100 | 100 | 100 | 100 | 100 |
| Batch | 2048 | 2048 | 2048 | 2048 | 2048 | 2048 |
| NCR Starting Epoch | NA | 30 | 30 | 30 | 30 | 30 |
| Hidden Layer (for NCR) | NA | 3 | 3 | 1 | 1 | 1 |
| NCR weight | NA | 0.3 | 0.5 | 0.8 | 0.8 | 0.8 |
| k | NA | 10 | 10 | 10 | 10 | 10 |

#### F.2 Mean Absolute Error

$$L_{NCR} := \frac{1}{m}\sum_{i=1}^{m} abs\left(\sigma(\mathbf{z_i}) - \sum_{j\epsilon NN_k} \frac{s_{i,j}}{\sum_k s_{i,k}}\sigma(\mathbf{z_j})\right) \tag{2}$$

Supplementary Table 11: AUROC, AUPRC, Sensitivity, and Specificity comparison between baseline and NCR models, across different amounts of error and test sets. In addition to label error in COVID-19 positive cases, there is also 0.5% label error in the negative controls. 0% error represents the original dataset, without any added label noise.

| Test Set | AUROC | | AUPRC | | Sensitivity | | Specificity | |
|---|---|---|---|---|---|---|---|---|
| | CE | CE+NCR | CE | CE+NCR | CE | CE+NCR | CE | CE+NCR |
| 0% error | | | | | | | | |
| PUH | 0.901(0.885-0.915) | **0.905(0.889-0.919)** | 0.568(0.521-0.614) | **0.580(0.539-0.625)** | 0.812(0.778-0.846) | **0.839(0.807-0.869)** | 0.839(0.832-0.846) | **0.844(0.837-0.852)** |
| UHB | 0.860(0.842-0.877) | 0.860(0.843-0.878) | **0.350(0.313-0.391)** | 0.344(0.311-0.386) | 0.818(0.787-0.848) | **0.820(0.789-0.850)** | 0.751(0.744-0.758) | **0.757(0.75-0.764)** |
| BH | 0.922(0.901-0.941) | **0.93(0.909-0.947)** | 0.706(0.638-0.765) | **0.738(0.682-0.800)** | **0.889(0.844-0.928)** | 0.882(0.833-0.924) | 0.777(0.755-0.799) | 0.777(0.758-0.799) |
| OUH "wave 2" | 0.875(0.867-0.883) | **0.883(0.875-0.891)** | 0.605(0.585-0.625) | **0.624(0.604-0.642)** | 0.834(0.819-0.848) | **0.847(0.833-0.860)** | **0.740(0.735-0.745)** | 0.734(0.729-0.739) |
| OUH "wave 1" | 0.846(0.831-0.860) | **0.859(0.845-0.872)** | 0.096(0.084-0.111) | **0.112(0.097-0.128)** | 0.792(0.766-0.817) | **0.812(0.787-0.837)** | **0.749(0.747-0.751)** | 0.742(0.740-0.744) |
| Error in 10% cases | | | | | | | | |
| PUH | 0.878(0.859-0.896) | **0.888(0.871-0.905)** | **0.559(0.512-0.606)** | 0.558(0.514-0.604) | 0.833(0.801-0.863) | **0.854(0.823-0.884)** | 0.716(0.707-0.724) | **0.74(0.732-0.749)** |
| UHB | 0.844(0.824-0.863) | **0.857(0.838-0.875)** | **0.328(0.292-0.372)** | 0.320(0.285-0.363) | 0.845(0.815-0.872) | **0.866(0.837-0.892)** | 0.659(0.652-0.667) | **0.660(0.652-0.668)** |
| BH | 0.924(0.902-0.943) | **0.932(0.912-0.949)** | **0.720(0.654-0.782)** | 0.694(0.632-0.767) | 0.917(0.876-0.951) | **0.938(0.901-0.970)** | **0.689(0.665-0.713)** | 0.684(0.661-0.709) |
| OUH "wave 2" | 0.861(0.852-0.870) | **0.872(0.863-0.880)** | **0.613(0.595-0.631)** | 0.589(0.570-0.610) | 0.851(0.838-0.863) | **0.881(0.869-0.893)** | **0.645(0.640-0.650)** | 0.613(0.608-0.618) |
| OUH "wave 1" | 0.822(0.807-0.838) | **0.849(0.835-0.863)** | **0.104(0.089-0.123)** | 0.101(0.088-0.120) | 0.833(0.810-0.856) | **0.863(0.842-0.885)** | **0.635(0.632-0.637)** | 0.615(0.612-0.617) |
| Error in 20% cases | | | | | | | | |
| PUH | 0.873(0.856-0.892) | **0.891(0.874-0.906)** | 0.503(0.457-0.553) | **0.557(0.514-0.601)** | **0.883(0.856-0.909)** | 0.880(0.851-0.907) | 0.579(0.569-0.588) | **0.645(0.636-0.655)** |
| UHB | 0.833(0.813-0.852) | **0.856(0.838-0.875)** | 0.313(0.278-0.356) | **0.366(0.328-0.411)** | 0.859(0.831-0.886) | **0.891(0.866-0.915)** | **0.593(0.584-0.601)** | 0.563(0.555-0.571) |
| BH | 0.922(0.903-0.94) | **0.928(0.907-0.946)** | 0.683(0.614-0.75) | **0.700(0.632-0.764)** | 0.938(0.901-0.969) | **0.965(0.939-0.987)** | **0.642(0.618-0.667)** | 0.614(0.590-0.638) |
| OUH "wave 2" | 0.851(0.842-0.86) | **0.869(0.86-0.877)** | 0.574(0.554-0.593) | **0.607(0.588-0.626)** | 0.891(0.879-0.902) | **0.916(0.905-0.926)** | **0.498(0.492-0.503)** | 0.492(0.487-0.498) |
| OUH "wave 1" | 0.816(0.799-0.832) | **0.834(0.819-0.849)** | 0.104(0.089-0.122) | **0.111(0.095-0.131)** | 0.862(0.839-0.882) | **0.887(0.868-0.906)** | 0.499(0.496-0.502) | **0.506(0.503-0.508)** |
| Error in 30% cases | | | | | | | | |
| PUH | 0.849(0.827-0.870) | **0.883(0.865-0.900)** | 0.474(0.428-0.520) | **0.502(0.457-0.553)** | 0.865(0.835-0.893) | **0.870(0.840-0.898)** | 0.543(0.533-0.553) | **0.640(0.631-0.649)** |
| UHB | 0.844(0.825-0.864) | **0.855(0.836-0.874)** | **0.330(0.294-0.375)** | 0.310(0.280-0.351) | 0.870(0.844-0.896) | **0.891(0.866-0.914)** | **0.587(0.579-0.596)** | 0.578(0.569-0.586) |
| BH | 0.911(0.885-0.932) | **0.928(0.906-0.948)** | 0.704(0.641-0.761) | **0.721(0.655-0.789)** | 0.938(0.902-0.967) | 0.938(0.901-0.969) | 0.592(0.568-0.616) | **0.668(0.643-0.692)** |
| OUH "wave 2" | 0.840(0.829-0.849) | **0.865(0.856-0.873)** | **0.552(0.531-0.572)** | 0.542(0.521-0.563) | 0.871(0.858-0.884) | **0.898(0.886-0.908)** | 0.512(0.506-0.517) | **0.520(0.514-0.525)** |
| OUH "wave 1" | 0.810(0.792-0.827) | **0.839(0.823-0.855)** | **0.112(0.097-0.131)** | 0.083(0.072-0.095) | 0.857(0.835-0.879) | **0.882(0.861-0.902)** | 0.480(0.477-0.482) | **0.521(0.518-0.524)** |
| Error in 40% cases | | | | | | | | |
| PUH | 0.857(0.836-0.879) | **0.880(0.862-0.898)** | 0.488(0.441-0.534) | **0.489(0.446-0.539)** | 0.883(0.856-0.908) | **0.911(0.887-0.936)** | 0.531(0.521-0.541) | **0.563(0.553-0.573)** |
| UHB | 0.836(0.816-0.856) | **0.853(0.833-0.871)** | 0.268(0.240-0.298) | **0.354(0.318-0.400)** | 0.886(0.861-0.910) | **0.900(0.874-0.922)** | 0.506(0.497-0.514) | **0.517(0.508-0.525)** |
| BH | 0.906(0.877-0.932) | **0.930(0.907-0.950)** | 0.640(0.578-0.703) | **0.703(0.637-0.765)** | 0.938(0.901-0.969) | **0.965(0.938-0.987)** | **0.541(0.517-0.568)** | 0.517(0.494-0.544) |
| OUH "wave 2" | 0.846(0.837-0.855) | **0.860(0.851-0.868)** | 0.477(0.459-0.494) | **0.547(0.526-0.568)** | 0.899(0.887-0.909) | **0.908(0.897-0.918)** | 0.456(0.451-0.462) | **0.465(0.459-0.471)** |
| OUH "wave 1" | 0.807(0.792-0.824) | **0.830(0.814-0.846)** | 0.052(0.046-0.057) | **0.098(0.085-0.114)** | 0.876(0.855-0.897) | **0.887(0.866-0.907)** | **0.453(0.451-0.456)** | 0.447(0.445-0.450) |

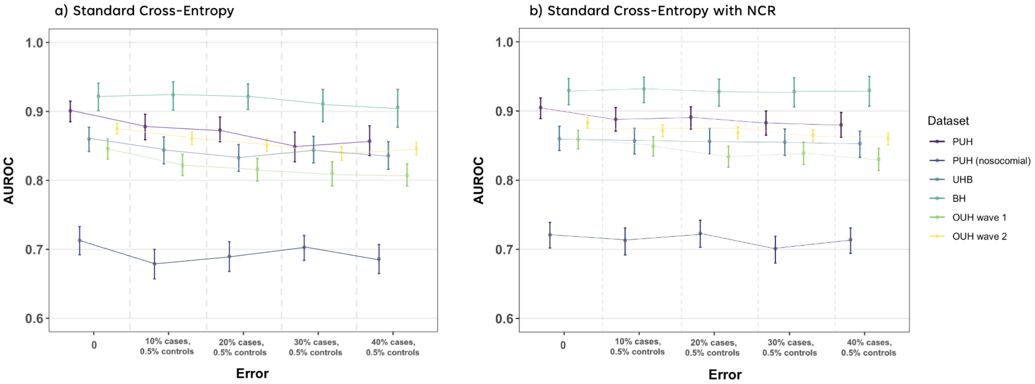


Supplementary Figure 6: Change in performance (AUROC) at different training error levels. Panel a) shows results when models are trained using standard cross-entropy, and panel b) shows results when models were trained with NCR.

Supplementary Table 12: Comparison of mean AUROC performance across different training set error levels.

| Test Set | CE | | CE + NCR | |
|---|---|---|---|---|
| | Mean | Std. | Mean | Std. |
| PUH | 0.872 | 0.020 | **0.889** | **0.010** |
| UHB | 0.843 | 0.010 | **0.856** | **0.003** |
| BH | 0.917 | 0.008 | **0.930** | **0.002** |
| OUH "wave 2" | 0.855 | 0.014 | **0.870** | **0.009** |
| OUH "wave 1" | 0.820 | 0.016 | **0.842** | **0.012** |

Supplementary Table 13: Hyperparameter values for final models (NCR term based on MAE) presented in main text.

| Loss Function | CE | CE + NCR | | | | |
|---|---|---|---|---|---|---|
| Error in Training Labels | 0% Error | 0% Error | 10% Error in Cases, 0.5% Error in Controls | 20% Error in Cases, 0.5% Error in Controls | 30% Error in Cases, 0.5% Error in Controls | 40% Error in Cases, 0.5% Error in Controls |
| Epochs | 100 | 100 | 100 | 100 | 100 | 100 |
| Batch | 2048 | 2048 | 2048 | 2048 | 2048 | 2048 |
| NCR Starting Epoch | NA | 30 | 30 | 30 | 30 | 30 |
| Hidden Layer (for NCR) | NA | 1 | 1 | 1 | 1 | 1 |
| NCR weight | NA | 0.05 | 0.03 | 0.03 | 0.04 | 0.03 |
| k | NA | 10 | 10 | 10 | 10 | 10 |

#### G Previous Studies Using Same COVID-19 Cohorts

Supplementary Table 14: Previously published COVID-19 status prediction results. using same datasets and patient cohorts. Sensitivity, specificity, and AUROC shown, alongside 95% confidence intervals, unless otherwise specified.

| Test Set | Sensitivity | Specificity | AUROC |
|---|---|---|---|
| **Soltan et al., 2022.** | | | |
| *Method: XGBoost + SMOTE + Threshold Adjustment (0.9)* | | | |
| OUH | 0.857 (SD 0.009) | 0.686 (SD 0.022) | 0.878 (SD 0.001) |
| PUH | 0.841 (0.825-0.857) | 0.713 (0.709 -0.718) | 0.872 (0.863 -0.882) |
| UHB | 0.788 (0.748-0.824) | 0.747 (0.738 -0.755) | 0.858 (0.838 -0.878) |
| BH | 0.743 (0.666-0.807) | 0.848 (0.825 0. 869) | 0.881 (0.851- 0.912) |
| **Yang et al., 2022.** | | | |
| *Method: Neural Network + SMOTE + Threshold Adjustment (0.85)* | | | |
| OUH | 0.844 (0.828-0.860) | 0.710 (0.704-0.717) | 0.777 (0.765-0.789) |
| PUH | 0.857 (0.842-0.873) | 0.672 (0.667-0.677) | 0.765 (0.752-0.777) |
| UHB | 0.847 (0.814-0.881) | 0.716 (0.708-0.725) | 0.782 (0.756-0.808) |
| BH | 0.847 (0.789-0.906) | 0.822 (0.799-0.845) | 0.835 (0.793-0.876) |
| **Yang et al., 2022.** | | | |
| *Method: Neural Network + Threshold Adjustment (0.85)* | | | |
| OUH | 0.762 (0.744-0.781) | 0.844 (0.839-0.849) | 0.878 (0.868-0.888) |
| PUH | 0.633 (0.585-0.681) | 0.903 (0.897-0.910) | 0.861 (0.837-0.885) |
| UHB | 0.714 (0.621-0.807) | 0.854 (0.839-0.870) | 0.878 (0.832-0.924) |
| BH | 0.724 (0.561-0.887) | 0.908 (0.869-0.948) | 0.880 (0.798-0.963) |

#### H Additional Case Studies

##### H.1 Data Availability

The eICU Collaborative Research Database is available online at `https://www.physionet.org/content/eicu-crd/2.0/`. The eICU Collaborative Research Database (eICU-CRD) is a publicly-available, anonymized database with pre-existing institutional review board (IRB) approval. The database is released under the Health Insurance Portability and Accountability Act (HIPAA) safe harbor provision. The re-identification risk was certified as meeting safe harbor standards by Privacert (Cambridge, MA) (HIPAA Certification no. 1031219-2).

The Adult (Census Income) Dataset is available online at `https://archive.ics.uci.edu/ml/datasets/Adult/`

##### H.2 Creating Noisy Labels

For ICU acute event prediction and income prediction, we randomly added incorrect labels at different noise ratios.

##### H.3 eICU Collaborative Research Database

Addressing the clinical applications of AI, the diagnosis of patients holds significant importance as it directly impacts clinical decision-making, allocation of resources, and healthcare expenditures. Thus, further analysis was performed using the eICU Collaborative Research Database (eICU-CRD) (Pollard et al., 2018) which is publicly available through PhysioNet (Goldberger et al., 2000).

In our experiments, we predict which acute condition might be developed by a patient during the course of an ICU stay, as defined through International Classification of Diseases, 9th Revision (ICD-9) codes. These are a system of alphanumeric codes used to classify and code diagnoses and procedures in medical billing and healthcare documentation. Previously, a similar undertaking involving both acute and chronic conditions was examined using the eICU-CRD dataset. In this study, 767 ICD-9 codes were grouped into 25 comprehensive diagnoses, which were then predicted using a BiLSTM model (Sheikhalishahi et al., 2020). Another study further grouped these diagnoses into their relevant systems and clinical specialties, before training a reinforcement learning model (Yang et al., 2022). Using consistent inclusion and exclusion criteria as these studies (detailed summary of criteria can be found in the Supplementary Material), we obtained three labels we aim to classify: acute cardiovascular event, acute respiratory event, and acute gastrointestional event. This grouping was selected to reflect clinic reality, where an emergency physician might consult with a system specialist to rule out a severe condition before admission to ICU, and to account for the relatedness of diagnoses within a system. For example, pneumonia is a leading cause of respiratory failure, and combining both diagnoses into a single "acute respiratory event" category reflects the systemic nature of the disease.

##### H.3.1 Inclusion and Exclusion Criteria

We selected adult patients (age > 18) with a minimum of 15 ICU records, grouped them into 1 hour windows, and asked our clinical team to reviewed the list of 25 diagnoses, removed 13 diagnoses considered chronic, non-acute, or poorly defined, and grouped the remaining 12 diagnoses into their relevant system and clinical specialties. We removed any samples that did not have a differentiable ICD9 code, or did not belong to any of the curated groups.

Supplementary Table 15: Acute event groups

| Label | Events |
|---|---|
| Acute cardiovascular event | Acute myocardial infarction, acute cerebrovascular disease |
| Acute respiratory event | Respiratory failure, insufficiency, arrest, pneumonia, pleurisy, pneumothorax, pulmonary collapse, other upper respiratory disease, other lower respiratory disease |
| Acute gastrointestinal event | Gastrointestinal hemorrhage |

Supplementary Table 16: Clinical predictors considered for predicting acute event diagnosis.

| Category | Features |
|---|---|
| Demographic features | Gender, age, ethnicity, height, weight |
| Measurements at hospital admission | Non-invasive systolic blood pressure, non-invasive diastolic blood pressure, non-invasive mean arterial pressure, heart rate, Supporting oxygen used at admission, blood oxygen saturation, Glasgow coma score, diagnosis at admission |
| Measurements at ICU admission | Glucose |

Supplementary Table 17: Summary of number of patients, COVID-19 positive cases, and ethnicity distribution for training, validation, and external test set cohorts included in the ethnicity debiasing task.

| | Training | Test |
|---|---|---|
| n, patients | 12,870 | 4,301 |
| Acute Cardiovascular | 5,214 | 1,776 |
| Acute Respiratory | 6,085 | 2,010 |
| Acute Gastrointestinal | 1,571 | 515 |

##### H.3.2 Acute Event Diagnosis Using NCR

To further evaluate the effectiveness of NCR, we demonstrate it's utility on an different task, using data from the eICU Collaborative Research Database (eICU-CRD) (Pollard et al., 2018; Goldberger et al., 2000). Our objective was to predict the occurrence of three acute events (cardiovascular, respiratory, gastrointestinal) during patients' stays in hospital intensive care units (ICUs). This task allowed us to evaluate the applicability of our model for a multi-class classification problem, which is commonly encountered in real-world applications.

In *Figure 6*, we observed that the model trained with NCR performs similarly to the model trained with standard cross entropy for cardiovascular and respiratory events. However, for gastrointestinal events, the model trained with NCR exhibits greater improvement in classification precision (p<0.001). This observation aligns with our expectations since gastrointestinal events have the lowest representation in the dataset, making this category more susceptible to the presence of noise. Consequently, NCR demonstrates its greatest positive impact on this specific group, effectively mitigating the model's tendency to overfit to noise. Full numerical values can be found in the Supplementary Material.

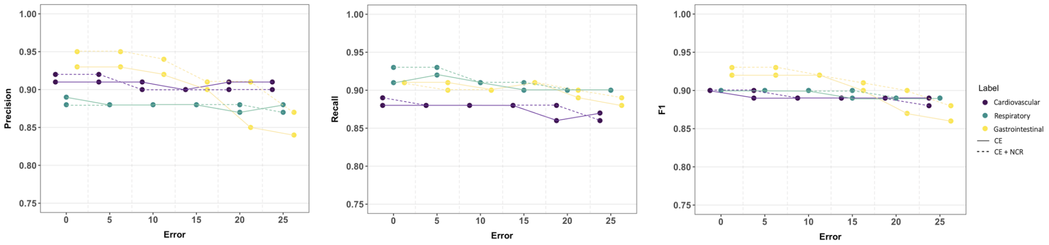


Supplementary Figure 7: Change in performance at different training error levels. Results presented as precision, recall, and F1 using a one-vs-rest method.

Supplementary Table 18: Comparison of mean AUROC (alongside standard deviation) performances across different training set error levels.

| Test Set | Precision | | Recall | | F1 | | p-value |
|---|---|---|---|---|---|---|---|
| | CE | CE+NCR | CE | CE+NCR | CE | CE+NCR | |
| 0% Error | | | | | | | |
| Cardiovascular | 0.91 | **0.92** | **0.89** | 0.88 | 0.9 | 0.9 | p<0.001 |
| Respiratory | **0.89** | 0.88 | 0.91 | **0.93** | 0.9 | 0.9 | p<0.001 |
| Gastrointestinal | 0.93 | **0.95** | 0.91 | **0.91** | 0.92 | **0.93** | p<0.001 |
| 5% Error | | | | | | | |
| Cardiovascular | 0.91 | **0.92** | 0.88 | 0.88 | 0.89 | **0.9** | p<0.001 |
| Respiratory | 0.88 | 0.88 | 0.92 | **0.93** | 0.9 | 0.9 | p<0.001 |
| Gastrointestinal | 0.93 | **0.95** | **0.91** | 0.9 | 0.92 | **0.93** | p<0.001 |
| 10% Error | | | | | | | |
| Cardiovascular | **0.91** | 0.9 | 0.88 | 0.88 | 0.89 | 0.89 | p<0.001 |
| Respiratory | 0.88 | 0.88 | 0.91 | 0.91 | 0.9 | 0.9 | p<0.001 |
| Gastrointestinal | 0.92 | **0.94** | 0.9 | 0.9 | 0.92 | 0.92 | p<0.001 |
| 15% Error | | | | | | | |
| Cardiovascular | 0.9 | 0.9 | 0.88 | 0.88 | **0.89** | 0.89 | p<0.001 |
| Respiratory | 0.88 | 0.88 | 0.9 | **0.91** | 0.89 | **0.9** | p<0.001 |
| Gastrointestinal | 0.9 | **0.91** | 0.91 | 0.91 | 0.9 | **0.91** | p<0.001 |
| 20% Error | | | | | | | |
| Cardiovascular | **0.91** | 0.9 | 0.86 | **0.88** | 0.89 | 0.89 | p<0.001 |
| Respiratory | 0.87 | **0.88** | 0.9 | 0.9 | 0.89 | 0.89 | p<0.001 |
| Gastrointestinal | 0.85 | **0.91** | 0.89 | **0.9** | 0.87 | **0.9** | p<0.001 |
| 25% Error | | | | | | | |
| Cardiovascular | **0.91** | 0.9 | **0.87** | 0.86 | **0.89** | 0.88 | p<0.001 |
| Respiratory | **0.88** | 0.87 | 0.9 | 0.9 | 0.89 | 0.89 | p<0.001 |
| Gastrointestinal | 0.84 | **0.87** | 0.88 | **0.89** | 0.86 | **0.88** | p<0.001 |

##### H.4 UCI Adult Dataset

The UCI Adult dataset is a widely used dataset in ML and data mining for classification tasks. It contains demographic and employment-related features of individuals, such as age, education level, marital status, occupation, and income, along with a binary label indicating whether the individual's income exceeds $50,000 per year.

###### H.4.1 Income Prediction Using NCR

We additionally evaluate NCR for income prediction, where the goal is to classify individuals into two income groups: those with income greater than $50,000 per year and those with income less than or equal to $50,000 per year. This data is also in tabular form, containing demographic and employment-related features of individuals, such as age, education level, marital status, occupation.

*Table 2* presents the AUROC and AUPRC scores achieved by models trained using standard cross entropy and models trained with the inclusion of NCR, considering different ratios of noisy labels. When compared to the standard baseline model, our approach significantly enhances performance, with improvements of up to 1.3% across all noise ratios ($p < 0.001$). Moreover, we demonstrate that utilizing NCR yields comparable, and slightly better, performance than standard cross entropy when there is no noise present ($p < 0.001$), further suggesting the general regularization effect of NCR. Additionally, we observe lower error rates at lower noise levels and a reduced improvement when NCR is incorporated (compared to the COVID-19 diagnosis task). This can be attributed to the utilization of a shallower neural network model for this prediction task, resulting in less error at lower noise levels and reduced susceptibility to overfitting to noise in general.

*Table 4* displays the mean AUROC and AUPRC (with standard deviation) obtained across different noise ratios. NCR consistently outperformed the baseline method, achieving the highest mean AUROCs on the held-out test set. Moreover, models trained with NCR exhibited lower standard deviations, indicating more consistent classification performance across various noise ratios. This suggests that the detrimental impact of increasing noise on performance was reduced compared to the baseline method.

Supplementary Table 19: AUROC and AUPRC comparison between baseline and NCR models, across different amounts of error. 0% error represents the original dataset, without any added label noise.

| Error (%) | AUROC | | AUPRC | | p |
|---|---|---|---|---|---|
| | CE | CE+NCR | CE | CE+NCR | |
| 0 | 0.896(0.890-0.903) | **0.898(0.891-0.904)** | 0.754(0.739-0.769) | **0.755(0.739-0.770)** | p<0.001 |
| 5 | **0.898(0.892-0.904)** | 0.897(0.891-0.903) | 0.751(0.736-0.766) | **0.756(0.741-0.771)** | p<0.001 |
| 10 | 0.894(0.887-0.900) | **0.898(0.892-0.904)** | 0.750(0.735-0.766) | **0.755(0.741-0.770)** | p<0.001 |
| 15 | 0.889(0.883-0.896) | **0.895(0.888-0.902)** | 0.743(0.728-0.759) | **0.758(0.744-0.774)** | p<0.001 |
| 30 | 0.887(0.880-0.893) | **0.889(0.882-0.895)** | 0.735(0.719-0.751) | **0.746(0.731-0.761)** | p<0.001 |
| 45 | 0.877(0.870-0.885) | **0.890(0.883-0.897)** | 0.728(0.712-0.744) | **0.745(0.729-0.761)** | p<0.001 |
| 60 | 0.870(0.862-0.877) | **0.883(0.876-0.890)** | 0.718(0.702-0.735) | **0.731(0.716-0.747)** | p<0.001 |

Supplementary Table 20: Comparison of mean AUROC and AUPRC (alongside standard deviation) performances across different training set error levels.

| Metric | CE | | CE + NCR | |
|---|---|---|---|---|
| | Mean | Std. | Mean | Std. |
| AUROC | 0.887 | 0.010 | **0.893** | **0.006** |
| AUPRC | 0.740 | 0.013 | **0.749** | **0.010** |

Supplementary Table 21: Hyperparameter values for final models (NCR term based on KL divergence) presented in main text.

| Loss Function | CE | CE + NCR | | | | | | |
|---|---|---|---|---|---|---|---|---|
| Error (%) in Training Labels | 0 | 0 | 5 | 10 | 15 | 30 | 45 | 60 |
| Epochs | 100 | 100 | 100 | 100 | 100 | 100 | 100 | 100 |
| Batch | 2048 | 2048 | 2048 | 2048 | 2048 | 2048 | 2048 | 2048 |
| NCR Starting Epoch | NA | 30 | 30 | 30 | 30 | 10 | 10 | 10 |
| Hidden Layer (for NCR) | NA | 1 | 1 | 1 | 1 | 1 | 1 | 1 |
| NCR weight | NA | 0.2 | 0.25 | 0.25 | 0.4 | 0.3 | 0.3 | 0.5 |
| k | NA | 10 | 10 | 10 | 10 | 10 | 10 | 10 |

### I Supplementary References


Goldberger, A. L., Amaral, L. A., Glass, L., Hausdorff, J. M., Ivanov, P. C., Mark, R. G., ... & Stanley, H. E. (2000). PhysioBank, PhysioToolkit, and PhysioNet: components of a new research resource for complex physiologic signals. circulation, 101(23), e215-e220.

Pollard, T. J., Johnson, A. E., Raffa, J. D., Celi, L. A., Mark, R. G., & Badawi, O. (2018). The eICU Collaborative Research Database, a freely available multi-center database for critical care research. Scientific data, 5(1), 1-13.

Sheikhalishahi, S., Balaraman, V., & Osmani, V. (2020). Benchmarking machine learning models on multi-centre eICU critical care dataset. Plos one, 15(7), e0235424.

Yang, J., El-Bouri, R., O'Donoghue, O., Lachapelle, A. S., Soltan, A. A., & Clifton, D. A. (2022). Deep Reinforcement Learning for Multi-class Imbalanced Training. arXiv preprint arXiv:2205.12070.
